# Pathologic subtyping of Alzheimer’s disease brain tissue reveals disease heterogeneity

**DOI:** 10.1101/2024.10.14.24315458

**Authors:** Tiffany G. Lam, Sophie K. Ross, Benjamin Ciener, Harrison Xiao, Delaney Flaherty, Annie J. Lee, Brittany N. Dugger, Hasini Reddy, Andrew F. Teich

**Author notes:** These authors share joint first authorship (Tiffany G. Lam, Sophie K. Ross). Corresponding author: Andrew Teich, Mailing address: 630 West 168^th^ Street, PH 15-124, NY NY, 10032.

## Abstract

In recent years, multiple groups have shown that what is currently thought of as "Alzheimer’s Disease" (AD) may be usefully viewed as several related disease subtypes. As these efforts have continued, a related issue is how common co-pathologies and ethnicity intersect with AD subtypes. The goal of this study was to use a dataset constituting 153 pathologic variables recorded on 666 AD brain autopsies to better define how co-pathologies and ethnicity relate to established AD subtypes. Pathologic clustering suggests 8 subtypes within this cohort, and further analysis reveals that the previously described continuum from hippocampal predominant to hippocampal sparing is well represented in our data. Small vessel disease is overall highest in a cluster with a low hippocampal/cortical tau ratio, and across all clusters small vessel disease segregates separately from Lewy body disease. Two AD clusters are identified with extensive Lewy bodies outside amygdala (one with a high hippocampal/cortical tau ratio and one with a low ratio), and we find an inverse relationship between cortical tau and Lewy body pathology across these two clusters. Finally, we find that brains from persons of Hispanic descent have significantly more AD pathology in multiple neuroanatomic areas. We find that Hispanic ethnicity is not uniformly distributed across clusters, and this is particularly pronounced in clusters with significant Lewy body pathology, where Hispanic donors are only found in a cluster with a low hippocampal/cortical tau ratio. In summary, our analysis of recorded pathologic data across two decades of banked brains reveals new relationships in the patterns of AD-related proteinopathy, co-pathology, and ethnicity, and highlights the utility of pathologic subtyping to classify AD pathology.

**Abbreviated summary:** Multiple groups have shown that what is currently thought of as "Alzheimer’s Disease" (AD) may be usefully viewed as several related disease subtypes. Here, we utilize two decades of banked cases to demonstrate how co-pathology and ethnicity intersect with one of the most widely used methods of pathologic subtyping.

## Introduction

Alzheimer’s disease (AD) is the leading cause of dementia in the elderly, and is a major cause of mortality and morbidity [1, 42]. As personalized medicine is pursued as a therapeutic strategy across multiple diseases [11, 36], an outstanding question in AD research is the extent to which AD may be usefully thought of as several related disease subtypes. AD pathology progresses in a stereotyped fashion, and tau and β-amyloid are both now recognized as spreading along established neuroanatomical stages [8, 49]. Although this pattern is generally true for the majority of cases, there is emerging evidence of significant variability in these trends, with tau deposition in particular showing different regional patterns. One of the foundational papers in this area identified AD subtypes using analysis of tau pathology in hippocampus and neocortex [35]. This analysis suggested that AD may segregate into three distinct pathologic subtypes – 1) typical AD, 2) hippocampal sparing (with relatively less tau pathology in hippocampus than typical AD and relatively more tau in neocortex than typical AD), and 3) hippocampal predominant (with these relationships reversed). Notably, these pathologic subtypes align with biological and demographic features, with hippocampal predominant subjects older than typical AD and more typically female, and hippocampal sparing subjects younger than typical AD, more typically male, and on average have a faster clinical progression than typical AD. These subtypes also relate to imaging metrics, as well as differential responses to therapy [13, 23, 53]. More recent work suggests that rather than bin AD into hippocampal sparing/predominant subgroups based on arbitrary thresholds of the hippocampal to cortical tau ratio, it is more useful to view all AD cases on a continuum using this variable [28].

The idea that AD may reflect different disease subtypes has prompted other groups to subtype AD into new classification groups based on pathologic data [10], transcriptomic data [37], and multi-omic data [25]. Analogously to the original paper on hippocampal-based AD subtyping, the newer papers that have identified subtype patterning have also identified features that segregate with subtypes, such as portability of brain multi-omic AD subtypes into multi-omic blood data in an independent cohort [25], subtype validation in different mouse models as well as in a replication cohort [37], and prediction of pathologic cluster grouping using MMSE, CSF data, and ApoE and MAPT genotype [10]. Additional work has suggested alternate pathologic categorizations that better align with atypical AD clinical variants [40].

In addition to varying methods of disease subtyping, a related issue is how common co-pathologies and ethnicity intersect with AD subtypes. Here, we exploit 20 years of brains banked at the New York Brain Bank of Columbia University and we apply clustering analysis to 153 pathologic variables recorded on these brains. Using the subset of 666 brains with a primary pathologic diagnosis of AD, we uncover new relationships between AD pathology and both Lewy body disease and vascular disease, including an inverse relationship between cortical tau and Lewy bodies in patients with extensive Lewy body co-pathology. In an effort to link our analysis to the prior literature in this area, we demonstrate that our clustering aligns with previously reported hippocampal sparing and hippocampal predominant phenotypes, and we further demonstrate that Hispanic persons are concentrated in specific AD subtypes and overall have more severe AD pathology across multiple regions. In summary, this work takes advantage of a large and well-characterized dataset to better define how co-pathology and ethnicity interact with AD pathologic subtypes, and builds on prior work documenting disparities in pathologic disease burden in persons of Hispanic descent.

## Materials and Methods

### Description of the New York Brain Bank

This study used all available cases with a completed neuropathology report from 2001 to 2022 at the New York Brain Bank (NYBB). The NYBB was founded in 2001 by Dr. Jean Paul Vonsattel (JPV). The NYBB serves as the pathology core for the Columbia University Alzheimer’s Disease Research Center and has additional prospective autopsy programs for Parkinson’s disease, amyotrophic lateral sclerosis (ALS), multiple sclerosis, Huntington’s disease, and Essential Tremor. For diagnosis, blocks are taken from the following regions (Brodmann areas (BA) are listed where appropriate): superior frontal cortex (BA 8, 9), posterior frontal cortex (BA 4), parietal cortex (BA 1, 3, 5, 40), calcarine cortex (BA17, 18, 31), hippocampal formation at the level of the lateral geniculate body, caudate with putamen and nucleus accumbens, globus pallidus with putamen and claustrum, amygdala, thalamus with anterior nucleus, midbrain, upper pons, lower pons, medulla, cerebellum with dentate nucleus, temporal pole, cingulate gyrus, subthalamic nucleus, and anterior hippocampus with entorhinal region. These 18 regions form the core of our diagnostic slide set, and pathologic variables from these blocks were used in this study. A standard slide of luxol fast blue/hematoxylin and eosin is produced for every block. In addition, immunohistochemistry for phospho-tau (AT8 at 1:200 dilution; Thermo Fisher; Catalog # MN1020), β-amyloid (6E10 at 1:200 dilution; BioLegend; Catalog # 803003), α-synuclein (KM51 at 1:40 dilution; Leica; Catalog # NCL-L-ASYN), and TDP-43 (C-terminal rabbit polyclonal at 1:500 dilution; Proteintech; Catalog # 12892-1-AP) is performed on blocks where these pathologies are expected to occur [8, 26, 33, 34] (totaling 30 immunostains as our minimal up-front set), in addition to 5 Bielschowsky silver stains on the first 5 blocks listed above (four cortical blocks + hippocampus).

Note that the above dilutions and antibodies represent the current protocol for brains at our bank. This study is a retrospective study over the past 20 years, and all of our staining is done on CLIA certified machines at Columbia University Irving Medical Center (in the Ventana automated slide stainer, without manual antigen retrieval and visualized using the Ventana ultraView universal DAB detection kit (Tucson, AZ) as recommended by the manufacturer). We do not have records of how the antibody concentrations and protocols have changed over the last 20 years at the immunohistochemistry core, and as such it is not possible to control for this batch effect over time. A similar point can be made concerning evolving criteria for diagnostic categories, particularly Alzheimer’s disease [2, 34]. Here, we are determining whether historical data can yield useful insights in AD pathophysiology, with all of the accompanying caveats that pertain to longitudinal pathologic data (this weakness of our study further discussed in the Discussion section). In addition, TDP-43 immunostaining was started in our brain bank in 2015 and our standard stain set established in 2018. As a result TDP-43 staining is not done on a sufficient number of subjects for the below clustering analysis (also a weakness discussed in the Discussion section).

### Description of variables used in this study

All of the cases used in this report were signed out between 2001 and 2022 by JPV (although JPV should be an author on this manuscript due to his extensive efforts, he is retired and not accepting authorship on manuscripts, see acknowledgements). Cases signed out by JPV had up to 276 variables recorded to varying degrees, and we exclusively used cases signed out by JPV for this study due to the fact that this is the large majority of cases at NYBB as of this study, and also to eliminate interobserver variability in pathologic grading. Of these 276 variables, we first eliminated variables using thresholding for feature completeness. Only features with at least 60% completeness across all available cases in the brain bank were retained, resulting in 196 pathologic features. Further filtering was applied at the sample level, requiring each case we retained for analysis to have at least 80% of the values across the 196 chosen features (these thresholding values were chosen after visual inspection of feature abundance curves, with thresholds chosen where abundance plateaus). After imputing missing data (see below), we then eliminated redundant features and features that represented summary statistics of other features, resulting in 153 unique pathologic features for clustering analysis. Additionally, pediatric cases and cases with unusual/rare diagnoses that did not fall into one of the diagnostic groups in Figure 1A were eliminated, which resulted in 1433 cases for subsequent clustering analyses. For the purposes of this study, “primary pathologic diagnosis” is defined by the top pathology diagnosis listed by JPV in the autopsy report, which he judged to be the diagnosis with the most severe pathology. Note however that there are frequently additional co-pathologies in most of our cases, as we are a brain bank for diseases of aging, where comorbidity is the rule rather than the exception [6, 26, 46]. Indeed, this varying co-pathology is a major focus of our analysis of AD cases in this paper. Of the 153 variables used for clustering analysis, 23 involve quantification of β-amyloid in various regions, 38 involve quantification of phospho-tau (tangles or neurites) in various regions, 36 involve quantification of atrophy or cell loss, 5 involve quantification of ischemia or vascular disease, 14 involve quantification of Lewy bodies, and 37 involve other miscellaneous categories (all variables with definitions listed in Supplemental Data). Pathologic data fell into three general categories; categorical (the pathologic feature is present/absent), subjectively graded (i.e. pathologic feature exists at low density, moderate density, or high density, based on experience), and precisely graded (i.e. each grade corresponding to a specific range of features such as tangles per 100x field). Examples of subjectively graded categories are displayed in Supplemental Figure 1.

**Figure 1:**
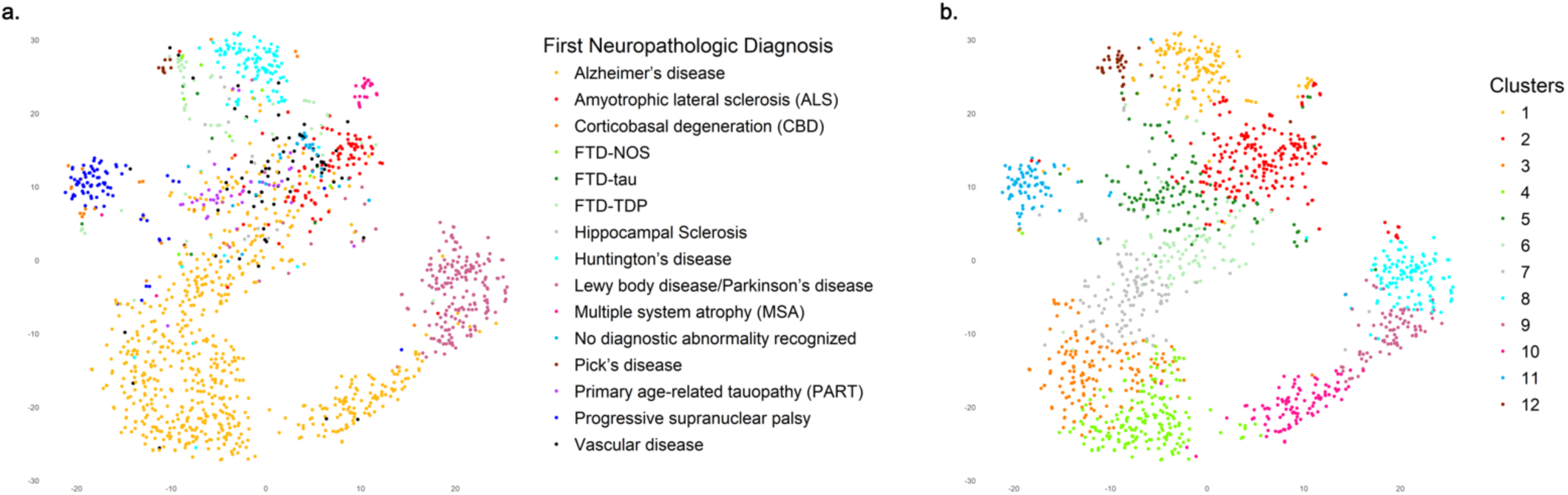
Clustering of 1433 cases across the NYBB identifies disease categories. (A) tSNE plot of 153 pathologic variables colored by primary pathologic diagnosis. (B) tSNE plot colored by the 12 identified clusters (see text for details).

### Clustering of pathologic variables

Missing data imputation was performed using the MICE (Multivariate Imputation by Chained Equations) method, which is particularly effective for handling missing data in datasets with mixed variable types. MICE generates multiple imputations to account for uncertainty and provides robust estimates by leveraging relationships between variables. This method was implemented using the mice package (version 3.16.0) in R [54]. To assess the dissimilarities between samples, a dissimilarity matrix was generated using the daisy function from the cluster package in R, applying the Gower metric to effectively handle the mixed dataset containing numerical and categorical pathologic variables.

The optimal number of clusters was determined using the gap statistic method, which compares the total within-cluster variation for different numbers of clusters against their expected values under a null reference distribution [50]. This was estimated with the clusGap function from the cluster package in R, utilizing the pam (Partitioning Around Medoids) algorithm as the clustering method, and analyzing up to a maximum of 20 clusters with 500 bootstrapping iterations. The optimal number of clusters was identified by examining the graph of gap statistic values, where the optimal point is where the gap statistic plateaus, indicating that additional clusters do not significantly improve the clustering structure. This can be defined a number of different ways [50], and we defined it as the minimum Gap(k) > Gap(k+1) – 2*SE(k+1), and also required that at least the next three successive Gap(k) values also pass this threshold to avoid local changes in the curve that are not reflective of the overall trend. For the full dataset, this method identified 12 clusters as optimal. When focusing on cases with a first neuropathological diagnosis of Alzheimer’s disease (AD), the same approach revealed that 8 clusters were optimal.

PAM was selected for its robustness to outliers and its use of actual data points as cluster centers (medoids). This characteristic enhances stability in the presence of noise, unlike centroid-based methods such as K-means, which can be significantly influenced by outliers [3]. By using real data points as medoids, PAM provides more meaningful cluster representatives compared to the abstract means used in K-means. This is particularly advantageous when dealing with noisy datasets, as K-means and hierarchical clustering methods can be sensitive to outliers, potentially leading to incorrect cluster assignments.

### Statistics

Non-parametric tests are used throughout this study (Spearman’s correlation coefficients, Mann-Whitney U test, Kruskal-Wallis ANOVA). Multiple hypothesis correction is done with false discovery rate, and all p-values have a threshold of 0.05. Age and sex were regressed from pathologic variables using the lm function in R. Hippocampal to cortical tau ratio was calculated by averaging the NFT values for CA1 and subiculum and dividing this by the average of the prefrontal cortex and parietal cortex NFT values for a given case.

## Results

The overall goal of this study is to use recorded pathologic data on brains in the New York Brain Bank (NYBB) to better define how co-pathology and ethnicity interact with AD pathologic subtypes. To do this, we took advantage of the fact that a majority of brains in the NYBB over the last 20 years have been signed out by a single neuropathologist (Dr. Jean Paul Vonsattel; although Dr. Vonsattel should be an author on this manuscript due to his extensive efforts, he is retired and not accepting authorship on manuscripts, see acknowledgements). We only used brains signed out by this pathologist for this study, which eliminates interobserver variability.

### Clustering of Pathologic Variables Reproduces Diagnostic Categories and reveals AD heterogeneity

We began by assembling all of the available recorded pathologic data on a cohort of 1433 cases regardless of diagnosis, and after thresholding based on data completeness (see Methods), we used a list of 153 variables for clustering (see Supplemental Data for list of variables). We first asked how useful this data was for recovering known diagnostic categories. To do this and to take advantage of the heterogeneous nature of the different pathologic feature categories, we performed Partitioning Around Medoids (PAM) clustering to identify pathologic clusters within the full dataset of 1433 brains, regardless of primary pathologic diagnosis. This analysis identified 12 optimal clusters (Figure 1B). When compared to the same datapoints labeled by primary pathologic diagnosis (Figure 1A), it is clear that several diagnostic categories correspond well to clusters, while others (such as MSA) are composed of multiple clusters. Notably, the majority of the AD cases are subdivided into six clusters, suggesting heterogeneity in this diagnostic group. Although tSNE is not itself a clustering technique, the fact that MSA cases are viewed separately in the tSNE plot suggests that there is information in our dataset that can identify these cases, even though the PAM algorithm preferentially splits AD into subcategories rather than assign a cluster to MSA. This further supports the view that there is significant heterogeneity in our AD cases that may be further explored.

To better define disease heterogeneity within AD we re-clustered all 666 AD cases separately, yielding 8 AD clusters (Table 1). To identify which pathologic variables varied the most by cluster, we performed Kruskal-Wallis ANOVA on each variable across all clusters and ranked by p-value (see Table 2 for select variables; Supplemental Data for full analysis). The top 6 variables by ranked p-value are all Lewy body related pathologies across multiple regions, and 3 are displayed in Table 2 as an example. There is also significant variation of AD-related pathologic variables (mostly tangles, neuropil threads, and neuritic plaques in various regions), as well as significant variation in the severity of small vessel disease pathology (i.e. cribriform change/lacunes in basal ganglia and pons). This analysis suggests that both AD and non-AD co-pathology distinguishes AD clusters in our analysis.

**Table 1:** Demographic and pathologic characteristics of the Eight AD clusters.

|  | 1 | 2 | 3 | 4 | 5 | 6 | 7 | 8 | Total |
| --- | --- | --- | --- | --- | --- | --- | --- | --- | --- |
| Age | 83.52 | **77.56** | **80.83** | **79.82** | **88.78** | **89.62** | **79.18** | 85.17 | 83.19 |
| Percent Hispanic | 6.4 | 10.78 | 7 | 17.11 | 3.85 | 1.37 | 0 | 9.43 | 7.36 |
| Percent Female | 56.8 | 58.82 | 57 | 57.89 | 58.25 | 63.01 | 39.39 | 60.38 | 57.59 |
| Braak NFT Stage | 5.74 | 5.99 | 5.88 | 5.93 | 5.13 | 4.42 | 5.42 | 3.81 | 5.41 |
| Braak LB Stage | 0.33 | 0.79 | 0.5 | 5.24 | 0.49 | 0.48 | 5.55 | 0.45 | 1.3 |
| Hippocampal/Cortical Tau Ratio | 1.75 | **1.00** | **1.23** | **1.13** | **2.86** | **3.61** | **2.52** | 2.00 | 1.76 |
| Cases Per Cluster | 125 | 102 | 100 | 76 | 104 | 73 | 33 | 53 | 666 |

*NFT = Neurofibrillary Tangle, LB = Lewy Body*
*Clusters 2-7 have age and hippocampal/cortical tau ratios that are significantly different when compared to all other cases using Mann-Whitney U test (bolded numbers, p-value less than 0.05).*

**Table 2:** List of selected pathology variables that significantly vary across AD clusters using a Kruskal-Wallis ANOVA (see Supplementary Data for full list)

| Pathology Variable | p-value | p-adj |
| --- | --- | --- |
| LB Inferior Temporal | 5.21E-112 | 7.98E-110 |
| LB OTG | 5.11E-111 | 3.91E-109 |
| LB Cingulate | 1.04E-106 | 5.31E-105 |
| NL Parietal | 4.34E-101 | 9.49E-100 |
| NL OTG | 1.03E-100 | 1.98E-99 |
| NFT Parietal | 9.11E-100 | 1.07E-98 |
| NFT Inferior Temporal | 1.38E-97 | 1.24E-96 |
| NFT Prefrontal | 2.43E-96 | 1.95E-95 |
| Cribriform change/lacunes CN | 5.75E-14 | 7.72E-14 |
| Vascular Sclerosis | 1.77E-04 | 2.23E-04 |

*LB = Lewy Body, OTG = Occipitotemporal Gyrus, NL = Neuronal Loss*
*NFT = Neurofibrillary Tangle, CN = Caudate Nucleus*

In an effort to organize this information further, we began by determining how AD pathology is varying across cluster. Prior work by Murray et al. have identified three subtypes of AD (typical, hippocampal sparing, and hippocampal predominant) which have been validated in several other reports [13, 23, 35, 53], and we began by determining how well these categories organize the distribution of AD pathology in our clusters. Although originally conceived of as separate groups, more recent work has demonstrated that the ratio of hippocampal to cortical tau pathology is more usefully thought of as existing on a continuum [28]. Inspired by this recent literature, we determined how this ratio varies in our data. To begin, we plotted this ratio by age across all samples (similarly to [35] we only included cases with Braak NFT stage 4 or higher, see Methods). This demonstrated a distribution that is similar to prior reports [28], with low hippocampal/cortical tau ratio cases being the primary population for younger patients, and high hippocampal/tau ratio cases existing exclusively at higher age points, and an overall positive significant correlation (r = 0.39; p-value = 1.42 x 10^-21^; Figure 2). Similarly to prior reports, this trend also skews female at higher hippocampal/cortical ratios, with 61% of subjects female in the upper half of the distribution, and 54.6% subjects female in the lower half of the distribution (a marginally significant difference; p-value = 0.02 using binomial test, p-value = 0.15 using Chi-squared test).

**Figure 2:**
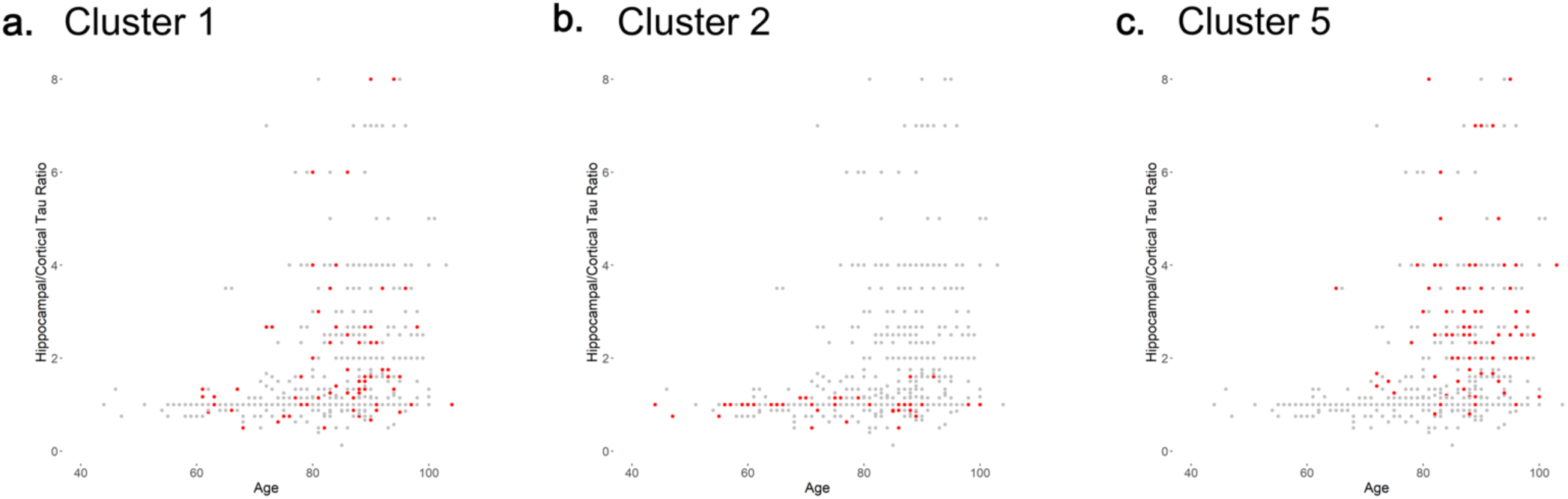
Hippocampal/cortical tau ratios for AD cases with Braak NFT stage of 4 or higher, plotted against age. In (A), (B), and (C), data points from representative clusters are shown in red against all other data points from AD cases. (A) Cluster 1, which is a cluster with an intermediate average hippocampal/cortical tau; (B) Cluster 2, which is a cluster with a lower average hippocampal/cortical tau ratio; (C) Cluster 5, which is a cluster with a higher average hippocampal/cortical tau ratio. Cluster numbers are taken from Table 1; see Supplemental Figure 2 for graphs of all clusters.

Next, we calculated the average hippocampal/cortical tau ratio value for each cluster (shown in Table 1), and also highlighted each cluster in our graph of ratio vs. age (Figure 2; see Supplemental Figure 2 for graphs for all 8 clusters). Both average ratios and cluster distributions reveal trends across samples. Cluster 1 has a ratio similar to the overall group (Table 1) and its samples are evenly distributed across ratio and age points (Figure 2), consistent with samples in this cluster being “typical” AD (i.e. without significant cortical or hippocampal skew). The next three clusters (2, 3, and 4) have lower than average ratio values (more similar to the hippocampal sparing phenotype), and the next three clusters (5, 6, and 7) have higher than average hippocampal/cortical ratio values (more similar to a hippocampal predominant phenotype). Cluster 8 is a cluster with low-AD pathologic change overall, and the ratio is less relevant for these samples, as the majority have a Braak NFT stage lower than 4. Note that the Murray et al. criteria, while well established, are not the only categorization scheme suggested for pathologic clustering, and a prominent alternative (Petersen et al.) has also been proposed based on hierarchical clustering of data [40]. It is more difficult to port categories based on data clustering across different datasets, and one advantage of algorithmically defined categories such as the Murray et al. criteria is the ease of applying these definitions to different datasets. Nevertheless, integration and cross-comparison of pathologic clustering schema should be the ultimate goal of these investigations (see Discussion).

### Lewy body pathology and vascular pathology segregate with clusters based on hippocampal/cortical tau ratio

Amongst the three low hippocampal/cortical tau ratio clusters (2, 3, and 4), cluster 2 has both the lowest average ratio and the lowest age of death amongst all clusters (77.56). The next two low ratio clusters have ratio values lower than average, although less extreme than cluster 2. Cluster 3 has the highest overall burden of small vessel disease pathology, and is higher than all other groups (Figure 3). Cluster 4 has extensive Lewy body disease pathology, which we compare to the other major Lewy body disease group (cluster 7) below.

**Figure 3:**
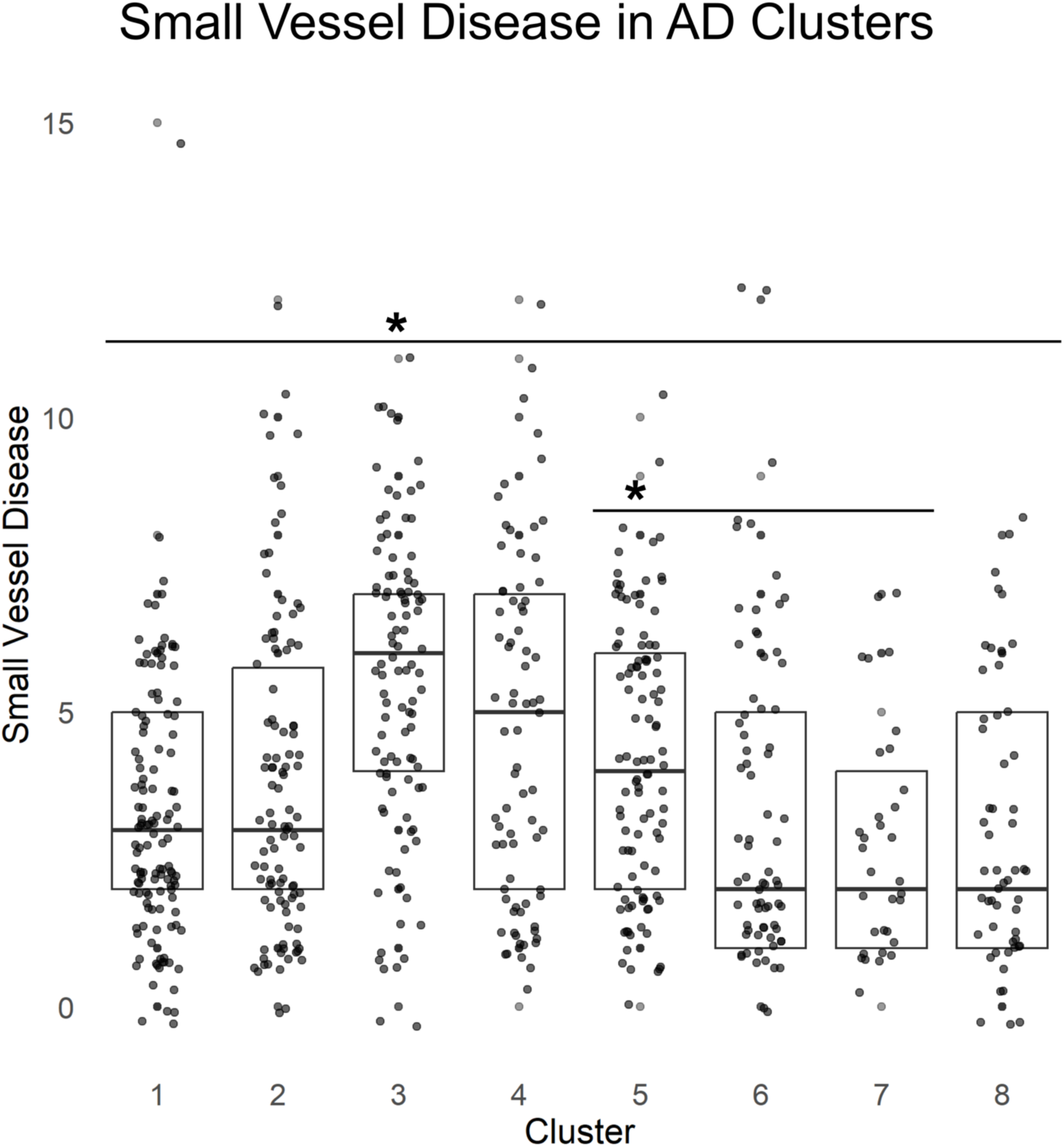
Small vessel disease scores (calculated as the sum of scores across four variables; Cribriform/Lacune pathology in pons, caudate, and putamen, as well as overall vascular sclerosis score; see Supplemental Data for definition of pathology variables), are shown for each AD cluster. Cluster 3 shows significantly higher pathology than all other clusters (Mann-Whitney U test p-value < 0.05 vs. cluster 4; less than 0.001 vs. all other clusters). Cluster 5 shows more pathology than the other two high hippocampal/cortical tau ratio clusters (clusters 6 and 7; Mann-Whitney U test p-value < 0.01 comparing 5 vs. 6 and 5 vs. 7).

Amongst the next three clusters with higher hippocampal/cortical tau ratios (5, 6, and 7), cluster 5 and 6 show demographic features consistent with the previously reported “hippocampal predominant” phenotype, with older ages of death and a higher % females in cluster 6. Cluster 6 has an even higher hippocampal/cortical ratio than cluster 5, and a higher age of death (the highest of all clusters, at 89.62). While clusters 5 and 6 are overall very similar, cluster 5 has a higher average Braak NFT stage and lower hippocampal/cortical ratio, and also significantly more small vessel disease (Figure 3), suggesting that small vessel disease is less severe in cases at the most extreme end of the hippocampal/cortical tau ratio.

The third high hippocampal/cortical ratio group (cluster 7) differs from clusters 5 and 6 in several aspects. The age of death (79.2) is a full decade younger than clusters 5 and 6, and the percent female (39%) is the lowest of all clusters, the opposite of the reported sex skew. Lewy body disease is extensive within this cluster, comparable to the Lewy body disease in cluster 4. In order to better understand how these two high Lewy body clusters relate to each other and the rest of the clusters, we examined Lewy body pathology across regions for these two clusters, and compared to cortical tau pathology across regions (Figure 4). Cases in cluster 4 (the low hippocampal/cortical tau ratio cluster) show extensive cortical tau pathology, while cluster 7 cases have minimal cortical tau (consistent with these cases having a higher ratio). Interestingly, Lewy body pathology shows the opposing trend; although present in both, there is comparatively less in cluster 4 and comparatively more in cluster 7. In an attempt to quantify this relationship, we analyzed correlation values of Lewy body and NFT/NPT scores across these cases. When analyzed across all AD cases, there was an expected positive trend, consistent with prior reports that Lewy body disease and tauopathy often synergize in AD [20, 22]. However, when only considering cases in these two subgroups with high Lewy body pathology, the opposing trend is seen, with significant negative correlations between extra-amygdala Lewy body pathology and cortical tau pathology in several areas. Note that Lewy body pathology across the neuraxis correlates inversely with cortical tau in these cases presumably because cortical tau strongly differentiates these cases with regards to overall AD pathologic burden; when expanded to NPT/NFT values across all areas, cortical tau continues to dominate the significant associations with Lewy body disease (Supplemental Figure 3). Also note that even in this sub-analysis the amygdala retains a positive trend between Lewy bodies and overall tau pathology, consistent with the unique role of the amygdala in AD-related Lewy body disease. In light of the above, one possible interpretation of cluster 7 is that cases with the most α-synuclein pathology are demographically more similar to PD (see Discussion for commentary).

**Figure 4:**
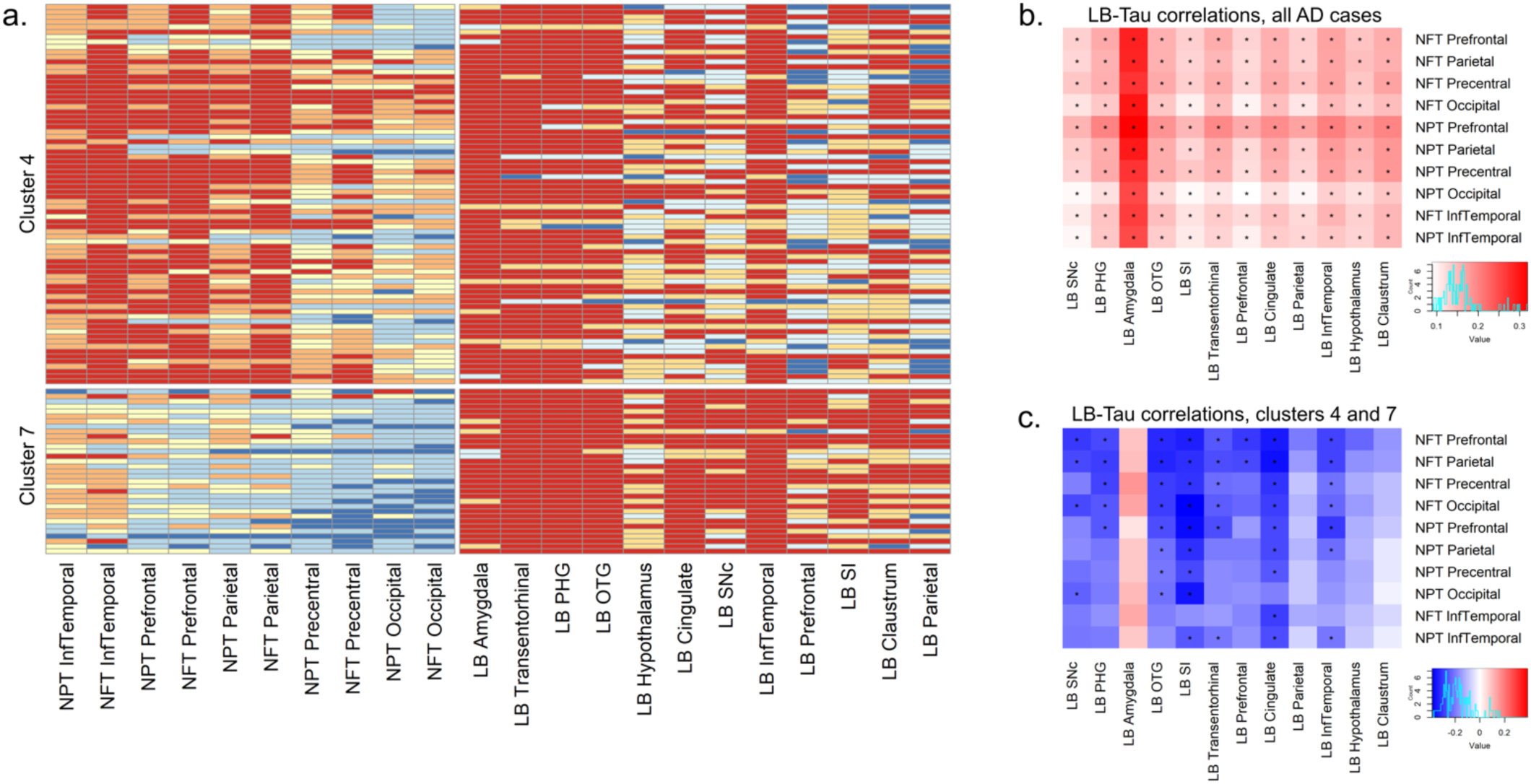
Lewy body pathology inversely covaries with tau pathology. (A) Tau pathology and Lewy body pathology heatmaps for AD clusters 4 and 7 (the two high Lewy body disease clusters in Table 1). Cortical tau burden (NFT and NPT) is high in cluster 4 and low in cluster 7. Lewy body pathology in contrast is more extensive in cluster 7 than in cluster 4. While tau and LB pathology co-correlate in the full AD cohort as expected (B), in the subset of patients in clusters 4 and 7 (C), the relationship is largely reversed (starred boxes = FDR adjusted p-value of Spearman’s correlation < 0.05). See text for details.

### AD pathology and associated co-pathology interact with Hispanic ethnicity

Finally, 49 of our AD subjects are of Hispanic heritage. Although we do not record country of origin for all subjects, approximately 70% of Hispanic residents from the Washington Heights community surrounding our institution are from the Dominican Republic [7], and so it is reasonable to assume that these demographics are reflected in the Hispanic persons in this study (also note that race was not included for all of these subjects, so we are only addressing Hispanic ethnicity here). We asked how all of the above pathologic metrics are affected by ethnicity from this perspective. The percent of persons who identified as Hispanic significantly varied across our AD clusters (chi-squared p-value across all groups = 0.003), with cluster 4 showing the highest percentage (for cluster 4 vs. all other cases, binomial test p-value = 0.0007; chi-squared p-value = 0.00126). It is particularly interesting that the Lewy body-related cluster 4 has the highest percentage of Hispanic cases, given the lack of Hispanic cases in the second Lewy body-related cluster 7 (see Discussion). In addition, Hispanic heritage predicted a higher overall burden of disease across multiple modalities, including neuronal loss, neuropil threads, neuritic plaques, neurofibrillary tangles, and Lewy body pathology (Figure 5). The age of Hispanic persons with AD was identical to non-Hispanic white persons with AD (mean of 82 in both cohorts), and there is an insignificant trend towards a higher percent of female subjects in Hispanic decedents (63.3%) in comparison to non-Hispanic white decedents (56.3%) (p-value = 0.2 by binomial test, p-value = 0.43 by Chi-square test). Consistent with this, regressing for age and sex has a negligible effect on the significance of these pathologic associations (Figure 5). In summary, these findings are overall consistent with prior work in this area [45], and suggest that ethnicity is significantly associated with the burden of AD and AD-related pathology (see Discussion).

**Figure 5:**
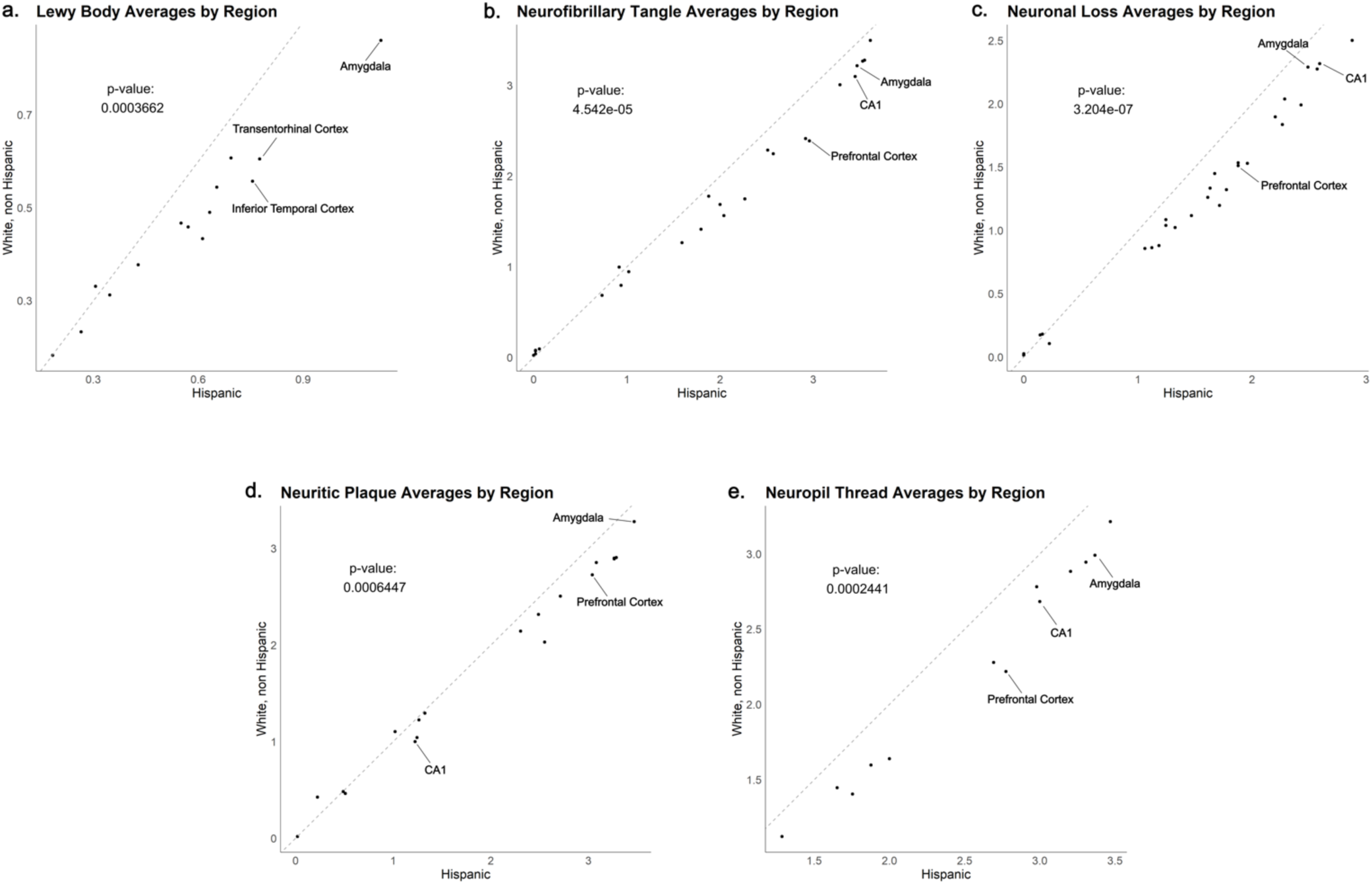
Hispanic persons with AD have more extensive pathology than non-Hispanic white persons with AD. For each graph, average values for a given pathology across all measured areas are plotted, with the average of Hispanic persons on the x-axis and the average of non-Hispanic white persons on the y-axis. Representative regions are labeled on each graph. A one-sample Wilcoxon test performed to determine whether the averages significantly deviate from a line with slope = 1 (which would indicate overall equivalent burden between groups) was performed on each graph, with p-value shown. While none of the individual values show a statistical difference between groups, an overall trend of more pathology in Hispanic persons is seen across all shown pathology categories. Note that after controlling for age and sex, all five variables retain significance (p-value < 0.001 for all variables).

## Discussion

AD subtyping is still a relatively new area, and the studies to date that use human tissue have approached the question from a variety of perspectives [10, 25, 37]. An additional limitation for this field has been that progress necessitates substantial effort and resources from an established brain bank with decades of well-characterized cases, which unfortunately limits scientific output in this area. Major questions overlying these investigations include 1) What is the best way to subtype AD?, and 2) How relevant are these subtypes for understanding AD pathogenesis and ultimately driving therapeutic strategy?

Here, we utilize two decades of banked cases at the NYBB to demonstrate how co-pathology and ethnicity intersect with one of the most widely used methods of pathologic subtyping. In light of this, it should also be noted that there are a number of alternative ways of subclassifying AD not evaluated in the current study. Uncommon clinical variants of AD (such as PPA) have also been linked to different patterns of tau pathology [19, 43, 56], and recent work from Petersen and colleagues have identified alternate clustering groups that align with these subtypes better than the hippocampal sparing/hippocampal predominant dichotomy of Murray et al. [40]. Specifically, Peterson et al. used hierarchical clustering to identify three subtypes with varying NFT burden (cortical predominant, high overall, and low overall), and demonstrated that cortical predominant cases have a high proportion of atypical clinical variants. In addition, Peterson et al. showed that their subgroups align with several clinical and cognitive measures better than the hippocampal sparing/hippocampal predominant types. As noted earlier, the ratio of hippocampal/cortical tau may be more useful than the original thresholds for hippocampal sparing/hippocampal predominant, particularly when looking for significant effects with lower sample sizes, and the hippocampal sparing/hippocampal predominant subtypes have aligned with additional imaging and clinical data in independent studies [13, 23, 53]. One advantage of the algorithmic definition of Murray et al. is the ease of analyzing any dataset for these trends, whereas groupings based on clustered data are often more difficult to port to other datasets, and we compare our data here to the Murray et al. definitions for both this reason and the literature supporting the relevance of these findings. Ultimately it will be important to achieve consensus on the best way to define AD subtypes, which should incorporate not just the Murray et al. criteria but additional perspectives such as the Peterson et al. categorization.

One of the most interesting findings in our data is the relationship of Lewy body pathology to AD neuropathologic change. LBD is found in 33% to 50% of persons with AD [12, 15, 55], and both AD and LBD share several common genetic risk factors [4, 12]. β-amyloid and tau have both been shown to interact with α-synuclein to mutually enhance aggregation [20, 22, 31], and Lewy bodies localized to the amygdala are so common in AD that a diagnostic category of “amygdala predominant Lewy body disease” was created to recognize this [32, 33]. Although the amygdala is an early incubator of Lewy body pathology in AD [32, 33, 41, 52], evidence suggests that Lewy bodies need to spread outside of this area to have a clinical effect [52]. Indeed, there is even some evidence that AD patients with amygdala-confined Lewy bodies may have α-synuclein aggregates that are less prone to propagation and are therefore less prone to cause disease [48]. Relatedly, Lewy body disease in the setting of AD appears to have a different pattern of spread from typical Parkinson’s disease. Specifically, “amygdala only LBD” is almost exclusively seen in the setting of AD [51, 52], and Lewy bodies are far more likely to spread in an amygdala-centric fashion in AD, bypassing a more typical progression from brainstem to cortex [9, 51]. In this context, it is notable that cluster 4 in Figure 4 has more extensive AD pathology, while Lewy body disease appears to be high in amygdala and limbic structures, and less prominent in brainstem and cortex, while cluster 7 has less extensive AD pathology but more extensive Lewy body pathology, including brainstem Lewy bodies. Given these pathology distributions, these two clusters may represents a dichotomy between AD specific amygdala-driven LBD in cluster 4 vs. more typical PD/LBD in the setting of AD pathology in cluster 7. This latter cluster is the smaller of the two (33 cases in cluster 7 vs. 76 in cluster 4), and one could speculate that this smaller cluster simply represents PD/LBD that received a primary pathologic diagnosis of AD somewhat equivocally over LBD. Indeed, as noted earlier this group is demographically more similar to PD, with a lower proportion of females and relatively younger age over the other AD clusters with high hippocampal/cortical tau ratios. Given the differing relationship between AD and LBD pathology in these clusters, it would be interesting to investigate whether AD pathology has different effects (in magnitude or degree) in synergistically driving LBD pathology in these two clusters, and whether they align with previously identified clinical LBD subtypes [16].

Along similar lines, it is interesting that small vessel disease appears more prominent in the low hippocampal/cortical ratio clusters, with cluster 3 showing the highest level of small vessel disease. Although vascular pathology and AD pathology are frequently seen in older individuals, the literature is mixed on whether these two are statistically associated with one another [14, 24, 27, 30], and a recent meta-analysis suggested that AD pathology and vascular pathology are uncorrelated [39]. Here, we show that small vessel disease segregates into specific AD clusters, which suggests that AD pathologic subtyping may help clarify the relationship between these two pathologies. Vascular disease and Lewy body disease also segregate differently in our analysis, and this discordance is particularly pronounced in the high hippocampal/cortical ratio clusters (i.e. comparing cluster 5 vs. 7). Interestingly, the above mentioned meta-analysis found that Lewy body disease negatively correlated with atherosclerosis and lacunar infarctions, and these were the only negative associations found across all age-related pathologies [39]. Although our findings here are broadly consistent with a dissociation between these two co-pathologies, future work should replicate our results or help clarify a negative association.

Our work also adds to the growing literature documenting how ethnicity influences the development of AD pathology [17, 21, 29, 38, 44, 45, 47, 58, 59]. Interestingly, Hispanic persons are not uniformly distributed across AD subgroups, and are particularly concentrated in cluster 4 (the high Lewy body disease group with a low hippocampal/cortical tau ratio). This concentration is particularly striking given the lack of Hispanic persons in the other high Lewy body group (cluster 7), with a high hippocampal/cortical tau ratio. These findings suggest that the development of extensive Lewy body co-pathology in Hispanic AD subjects may mostly or exclusively occur in the setting of high cortical AD pathology, although our sample size is low and these results need to be replicated. We also found increased burden of several individual pathologic variables in Hispanic persons. Although some have found evidence of higher tau deposition in Hispanic persons that survives adjustment for age and sex [45], others have found higher tau burdens that do not survive adjustment for sex, age of onset, or disease duration [44]. Hispanic persons with AD have also been shown to have more extensive co-pathology [17, 45, 58, 59], including more small vessel disease [17, 59] (although see [45]), and more Lewy body disease [58] (although see [44]). We found a higher burden of several pathologic variables in Hispanic decedents, and in our data these associations survive adjustment for age and sex. As noted above, higher levels of AD pathology in Hispanic persons are variably dependent on different demographic factors [44, 45]. Future work should examine how these factors interact with ethnicity, and why associations with ethnicity variably survive controlling for demographic factors in different studies. An additional challenge when comparing persons who identify as Hispanic to persons who do not is potential differences in socioeconomic status, which are known to independently affect AD progression [57], and are rarely controlled for given the difficulty of routinely obtaining an objective measure of this information. Indeed, there are many aspects influencing differences in ethnicity and race reported in the literature, including but not limited to access to care, poverty, education, living conditions, culture, stress, and systemic, institutional, and individual racism [5, 45, 60, 61]. Finally, in addition to different ways of measuring pathology, the literature examining the role of Hispanic ethnicity on AD pathologic change needs to account for the diverse range of backgrounds that identify as Hispanic [18], as well as chronically low brain bank enrollment of Hispanic patients. Although we do not keep track of country of origin for all subjects, our local community in Washington Heights is predominantly of Caribbean Hispanic origin (from the Dominican Republic) [7], and it is reasonable to assume that the majority of our local Hispanic identifying donors reflect this background. Given all of these confounding issues, it is notable that increased AD pathology is still reproducibly found in Hispanic individuals in comparison to non-Hispanic white subjects. The work presented here adds to this literature, and underscores the importance of continuing to examine this phenomenon and understand its etiology.

Our work has several other limitations. This study is a retrospective study over the past 20 years, and as noted in the Methods, we do not have records of how the antibody concentrations and protocols have changed over the last 20 years at the immunohistochemistry core where our staining is performed. As such it is not possible to control for this batch effect over time. A similar point can be made concerning evolving criteria for diagnostic categories, particularly Alzheimer’s disease, which was updated in 2012 [2, 34]. Here, we are determining whether historical data can yield useful insights in AD pathophysiology, with all of the accompanying caveats that pertain to longitudinal pathologic data. Despite these caveats, the fact that we are able to reproduce aspects of previously defined pathologic subtypes is partial validation that our approach is scientifically useful. Finally, TDP-43 immunohistochemistry was started at the NYBB in 2015 and our final stain set with AD cases was established in 2018, and as such we do not have data on a high enough number of subjects in this study to allow for inclusion of TDP-43 data in our clustering. Future work will focus on staining historical cases from our cohort for TDP-43, which will allow for more complete analysis of the role of TDP-43 proteinopathy in defining disease subtypes in AD and other neurodegenerative diseases.

In conclusion, we have presented clustering data from cases from our brain bank that identifies and builds on previously established AD subtypes. We have shown that AD clustering identifies trends in the coincidence of comorbid disease, and have provided additional evidence for the role of ethnicity in the development of AD pathology. Future work should further integrate clustering schema across the field and work towards a consensus definition of optimal AD subtypes.

## Supplemental Data

This contains a list of all pathologic variables and definitions; also shown are results from a Kruskal-Wallis ANOVA test for all variables across AD groups (select variables from this list are displayed in Table 2).

## Abbreviations

**Pathology**
NP: neuritic plaque
NL: neuronal loss
NFT: neurofibrillary tangle
NPT: neuropil threads
LB: Lewy body
IP: immature plaque (diffuse plaques)
GCI: glial cytoplasmic inclusion
BN: ballon neuron
Marinesco_B: Marinesco body

**Regions**
SNc: Substantia nigra pars compacta
CA1-4: CA1-4 region of hippocampus
OTG: occipitotemporal gyrus
PHG: parahippocampal gyrus
SI: substantia innominata
TempPole: temporal pole
SNr: Substantia nigra pars reticulata
RN: red nucleus
GPe: external globus pallidus
GPi: internal globus pallidus
CN: caudate nucleus
STN: sub-thalamic nucleus
WM: white matter
HF: hippocampal formation

## Ethics approval

This work uses deidentified pathologic variables and demographic data from autopsy subjects, and has been determined to be not human subject research by the Columbia University IRB.

## Data availability

The datasets analyzed during this study are available from the corresponding author on reasonable request.

## Acknowledgements

This work would not have been possible without the meticulous work of Dr. Jean Paul Vonsattel over a 20 year period. Dr. Vonsattel not only founded the New York Brain Bank, but generated all of the pathologic data used in this manuscript. As such, he should be an author, if not a co-senior author, on this manuscript, but he is currently retired and not accepting authorship on any manuscripts. At minimum, we would like to thank him for his work that made this manuscript possible, as well as his mentorship to all of the members of the New York Brain Bank. We dedicate this manuscript to him and his legacy at Columbia University.

