## Supplemental Figure for "Pathologic subtyping of Alzheimer’s disease brain tissue reveals disease heterogeneity"

**Supplemental Figure 1:** Examples are shown of semi-quantitative variables.

### Neuropil Threads

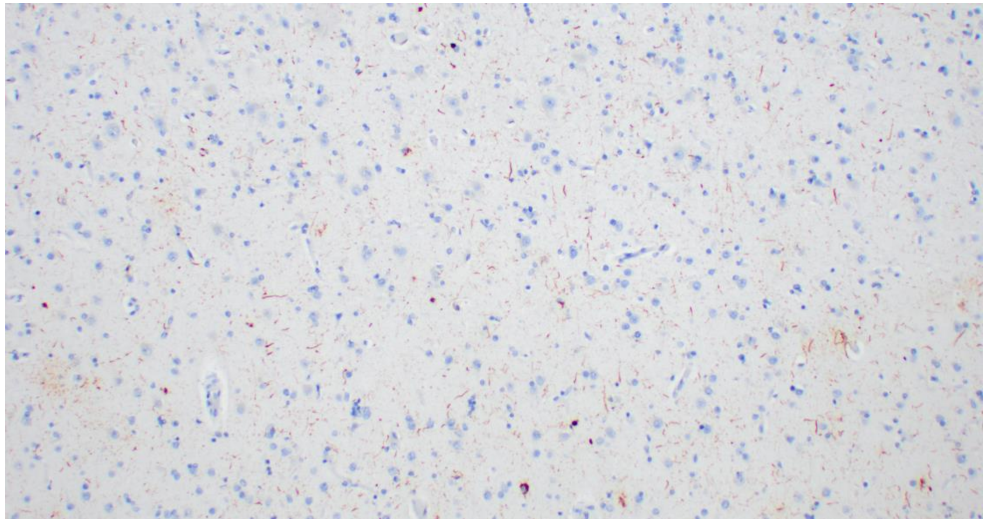

Grade 1

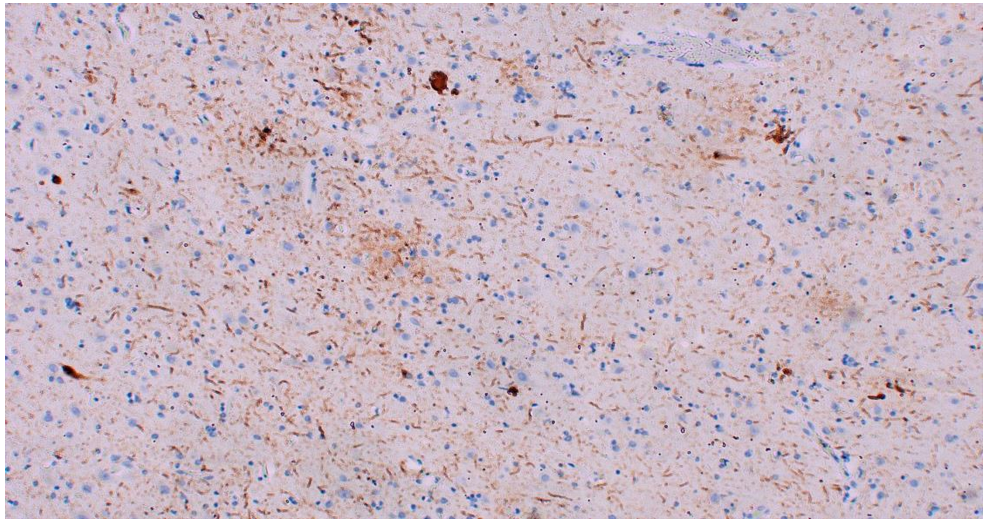

Grade 2

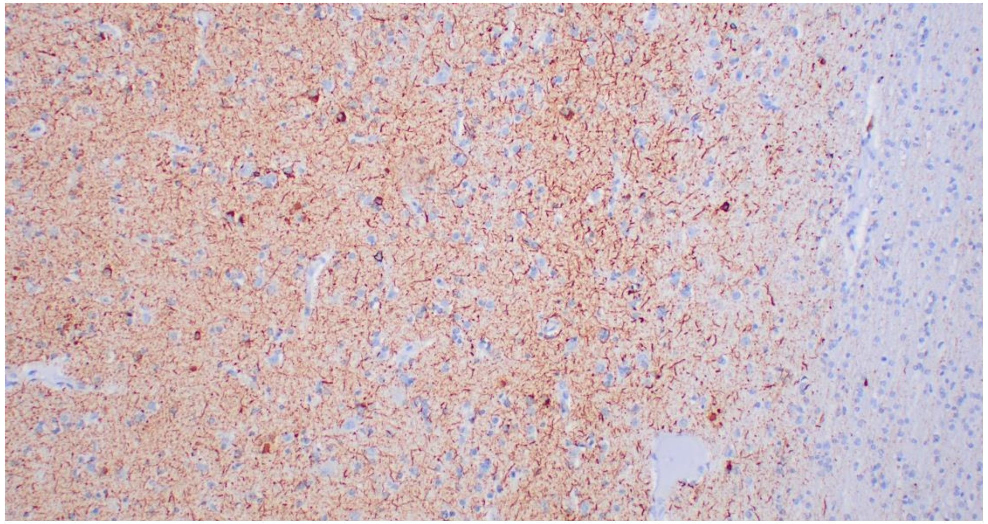

Grade 3

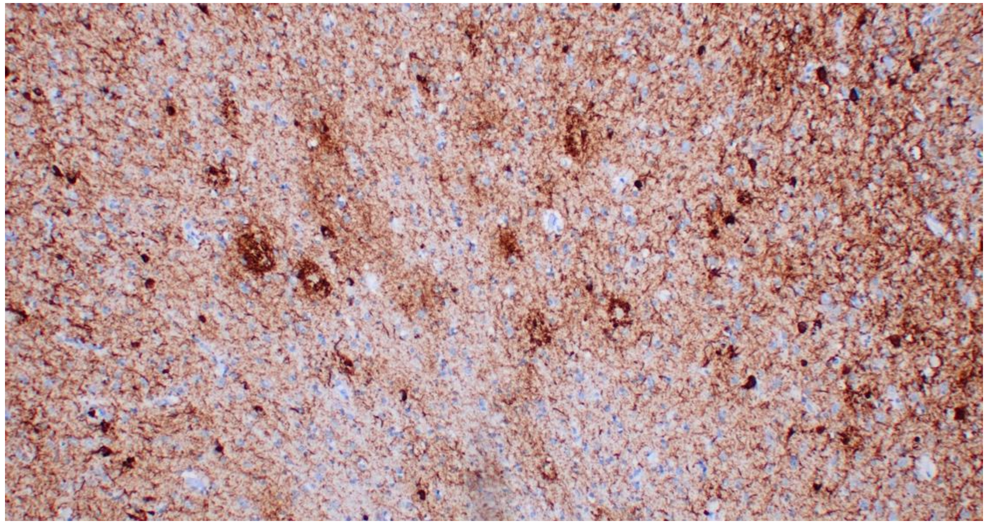

Grade 4

### Pyramid myelin loss

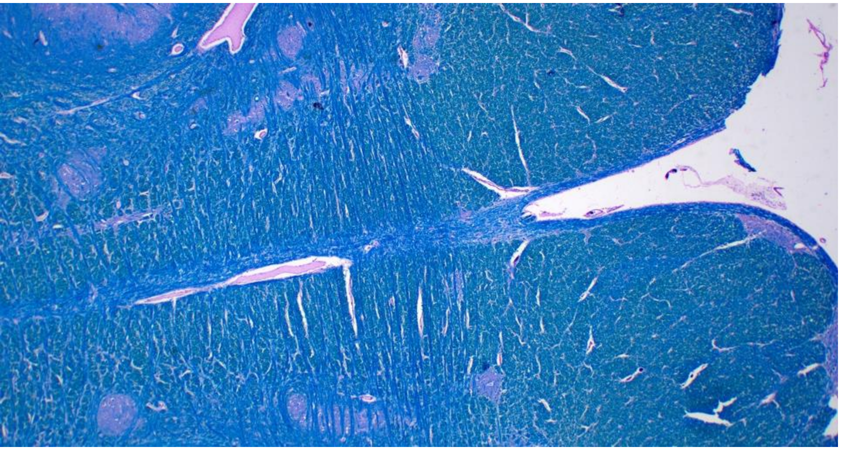

Normal

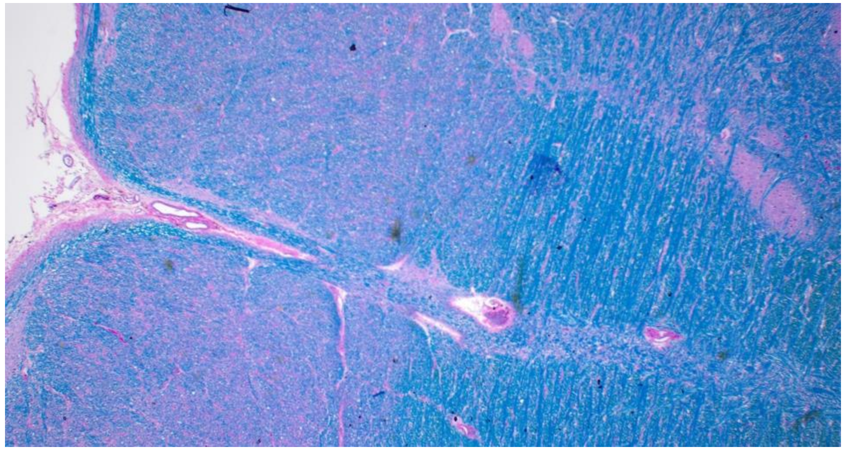

Loss 1

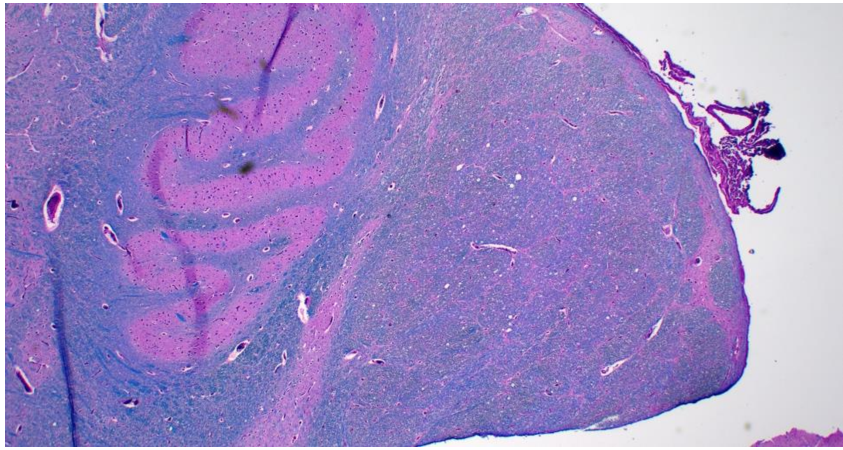

Loss 2

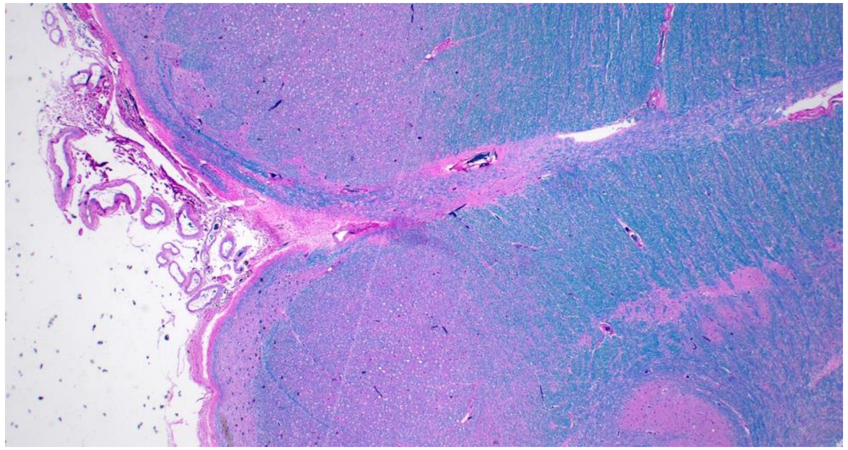

Loss 3

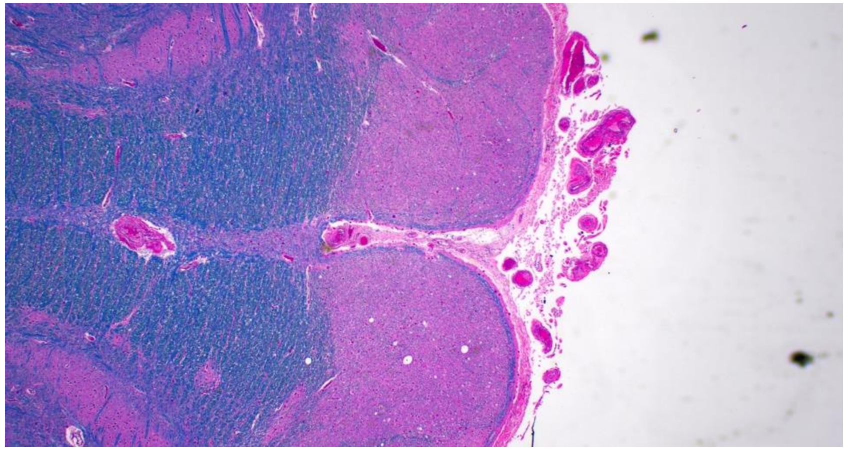

Loss 4

### Status Spongiosus

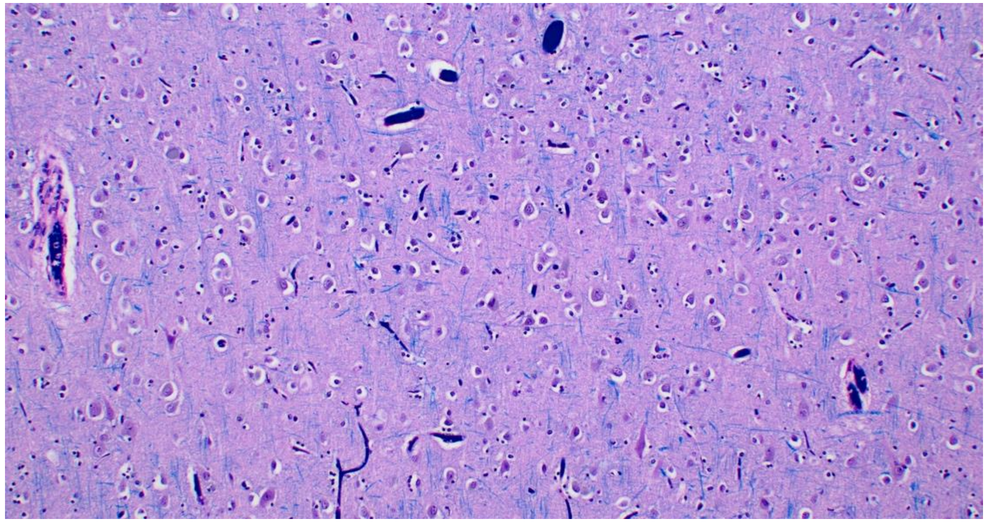

Grade 1

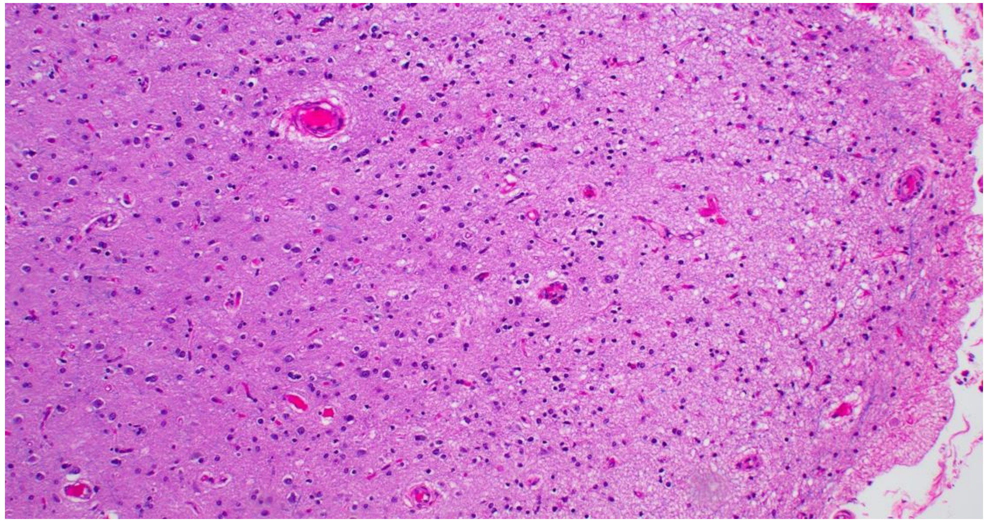

Grade 2

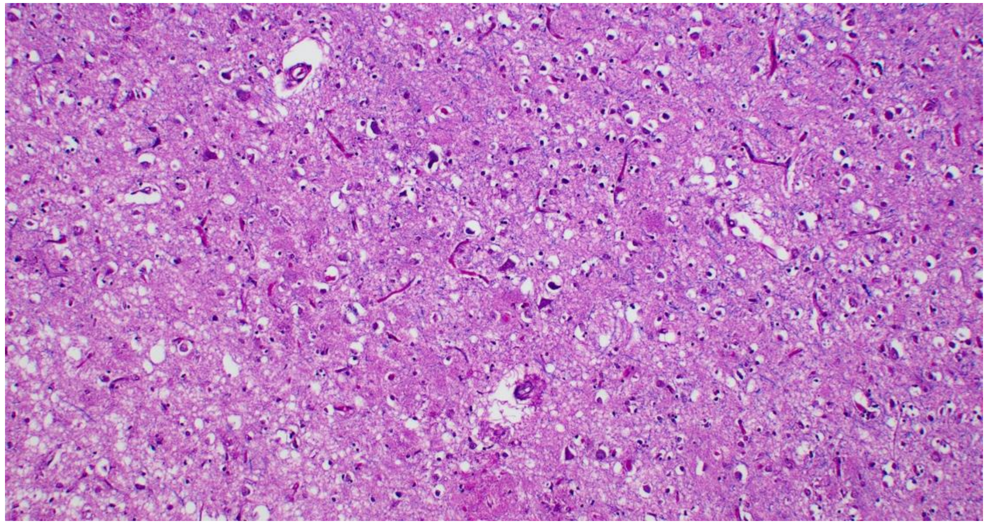

Grade 3

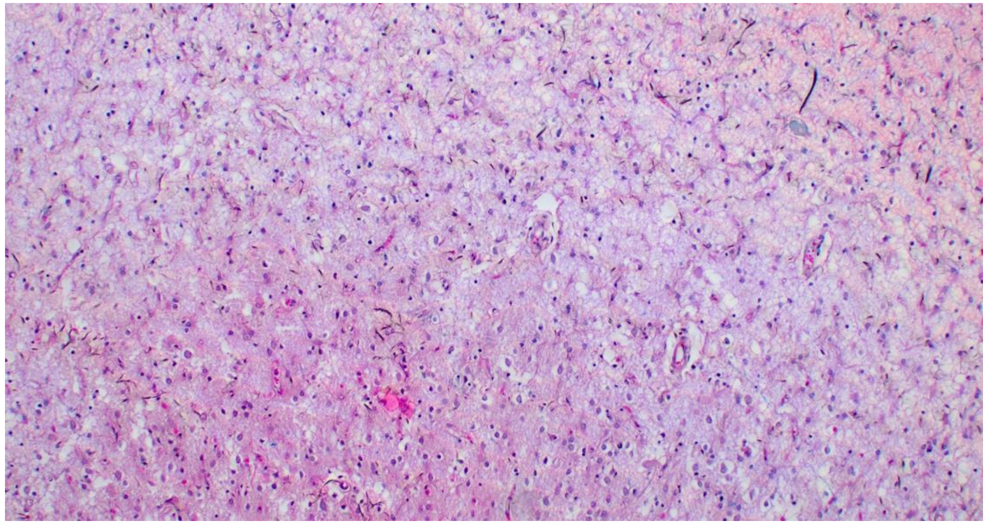

Grade 4

### Neuronal Loss

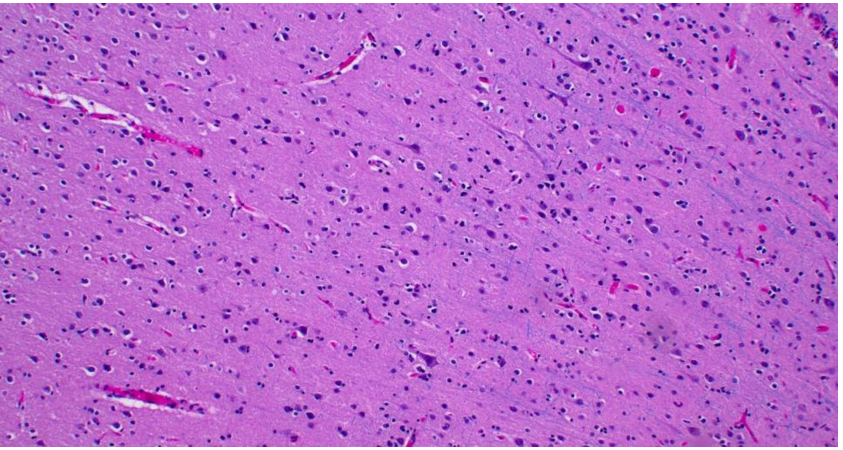

Normal

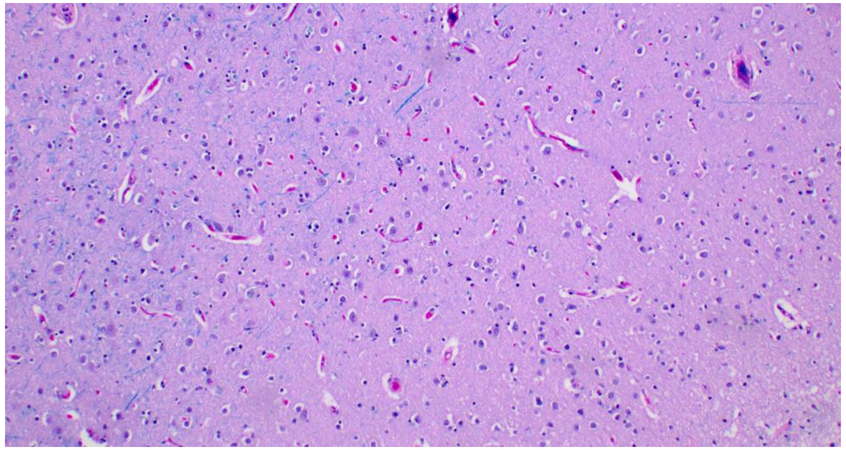

Grade 1

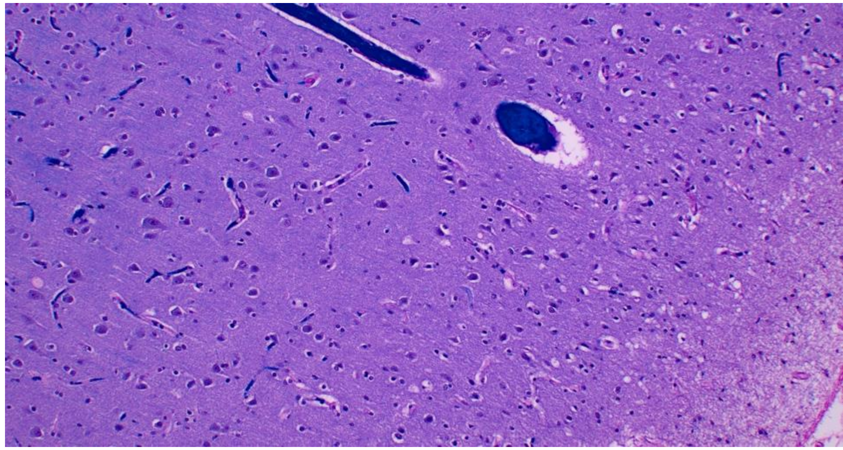

Grade 2

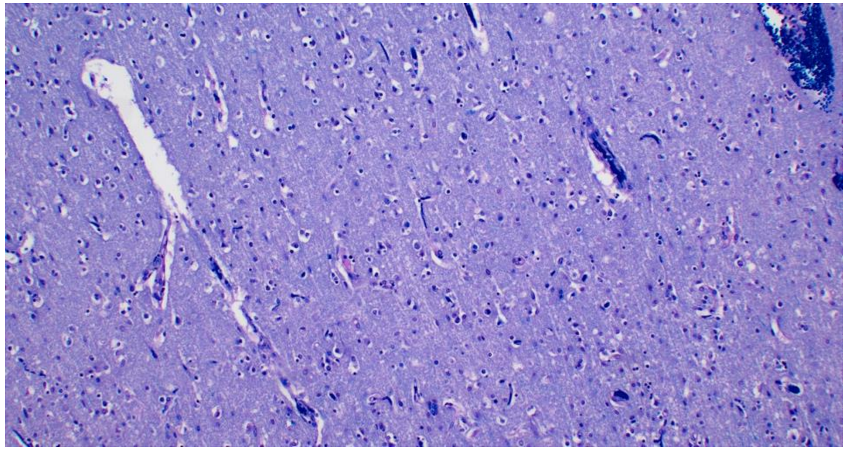

Grade 3

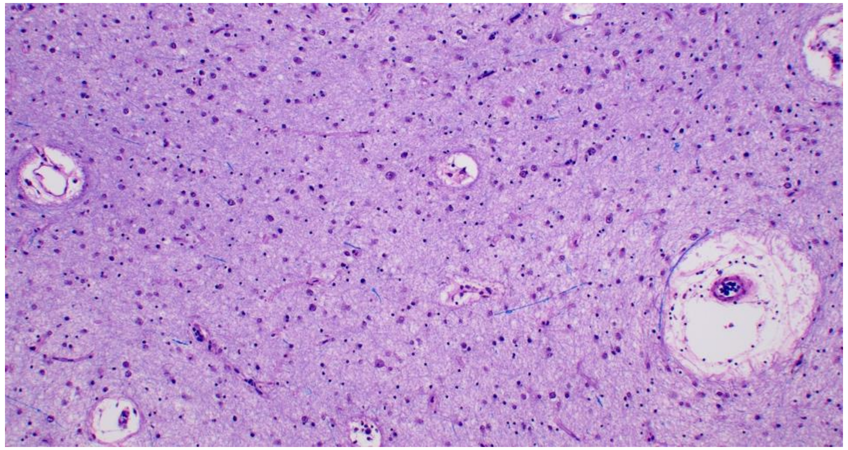

Grade 4

### Atrophy, Grade 0 (no atrophy)

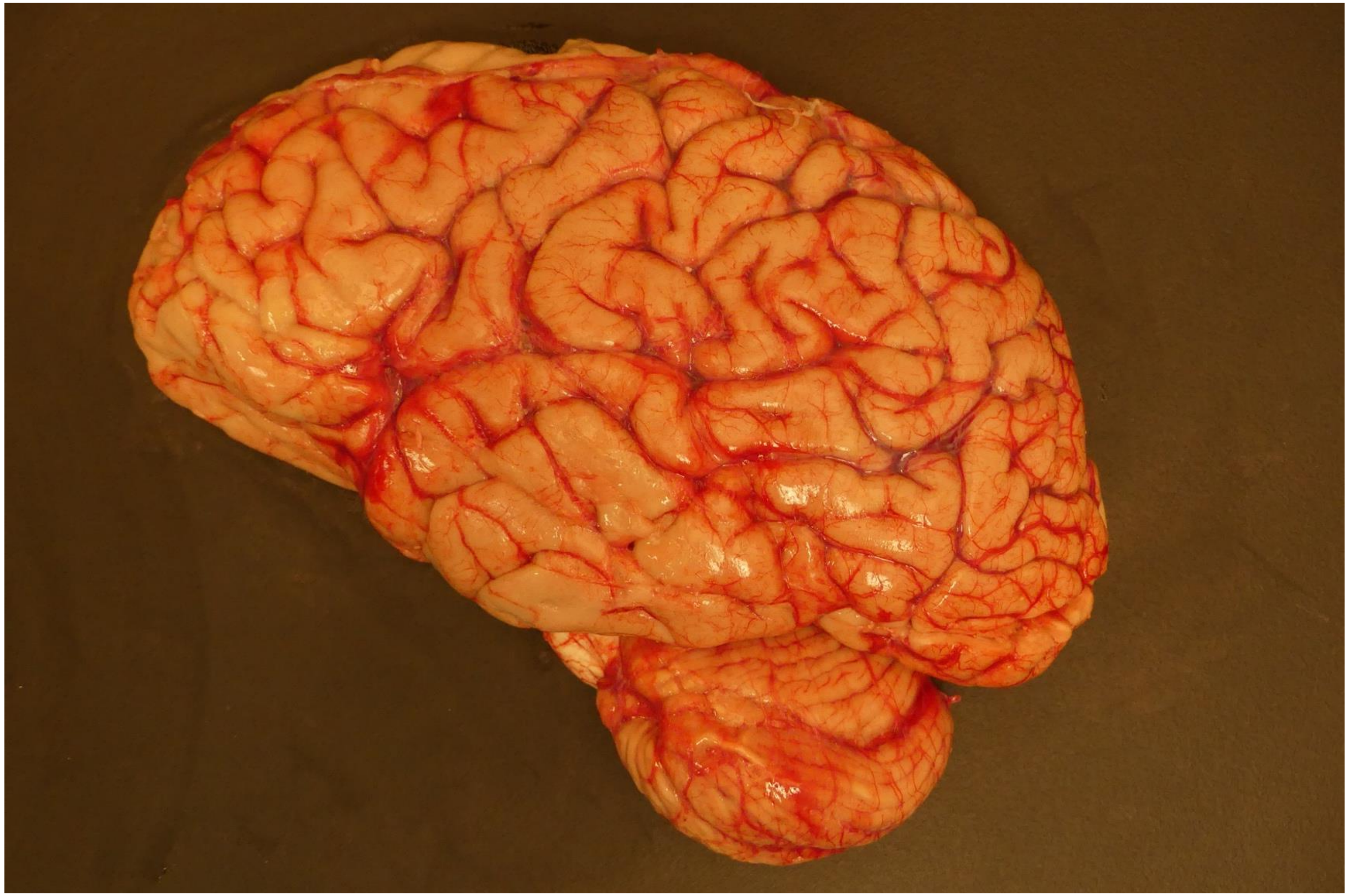

### Atrophy, Grade 1

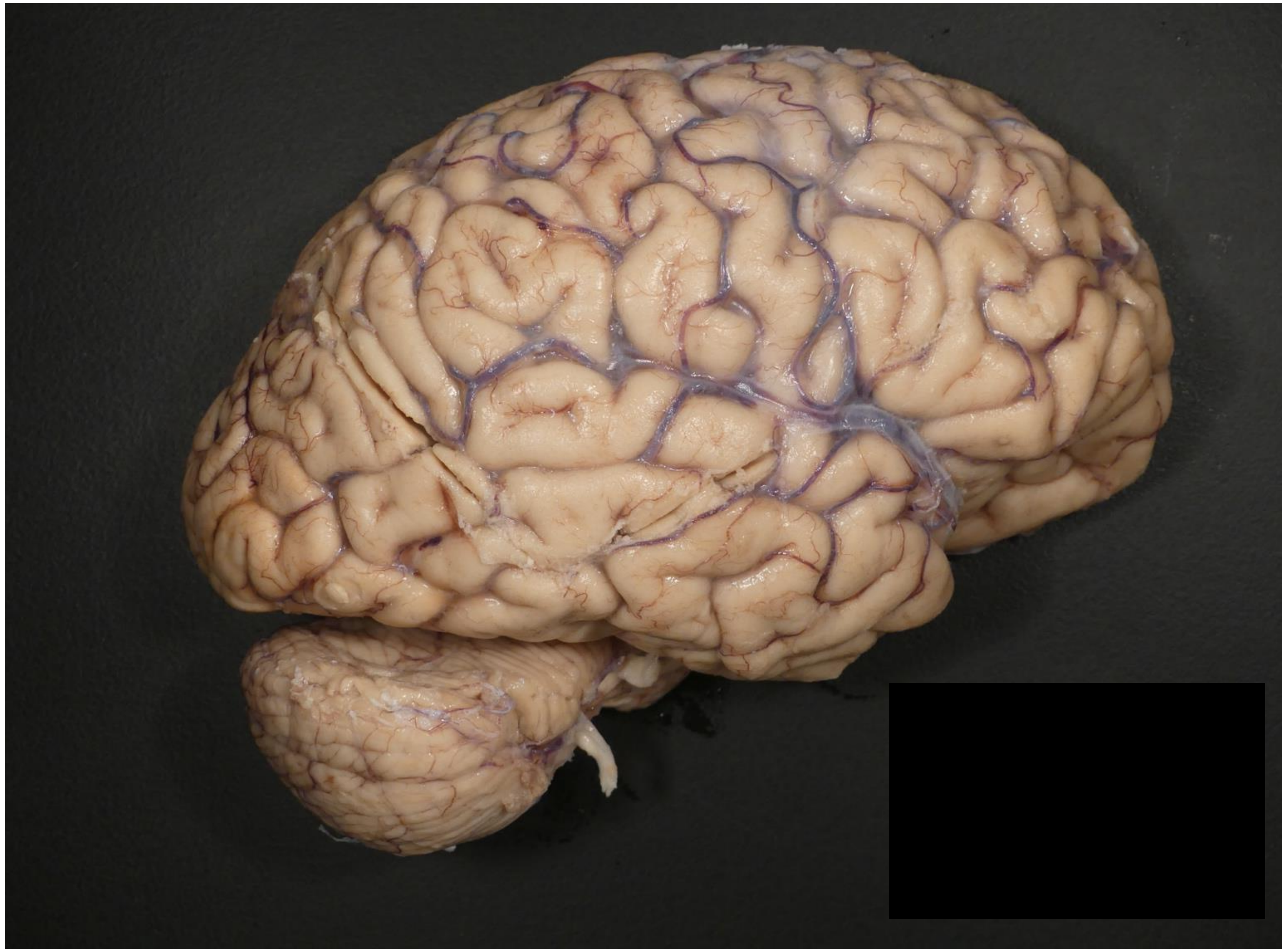

### Atrophy, Grade 2

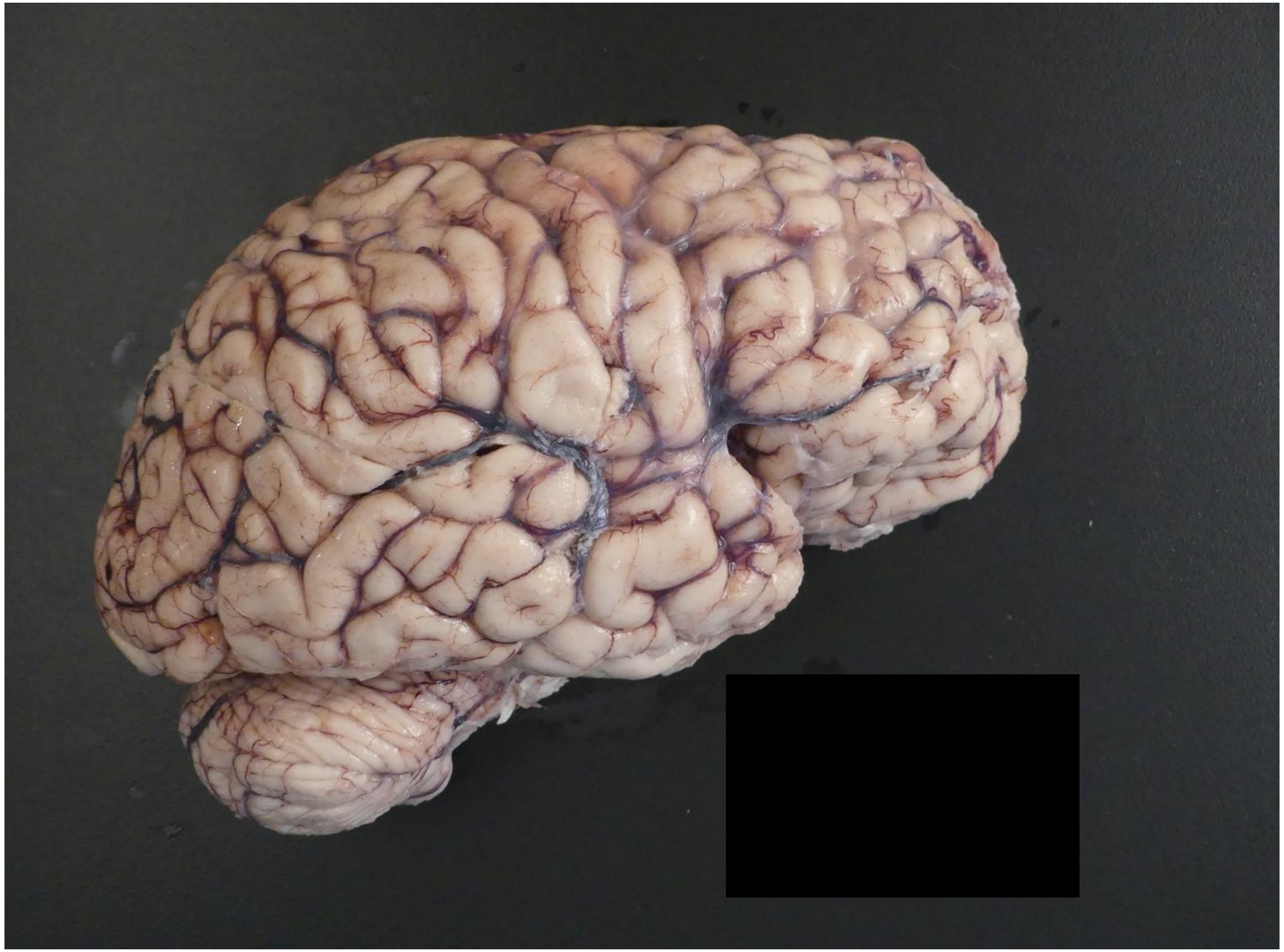

### Atrophy, Grade 3

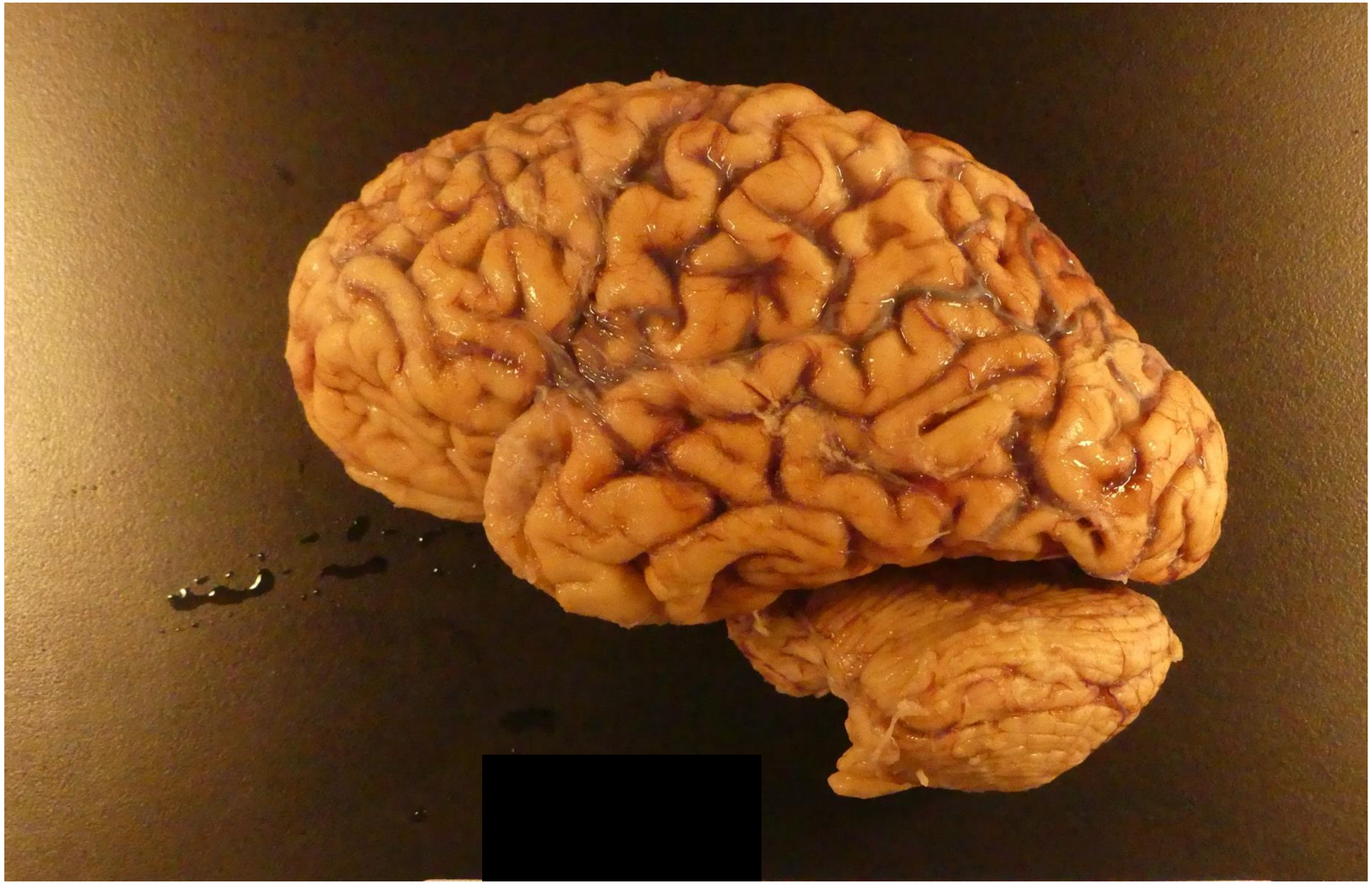

### Atrophy, Grade 4

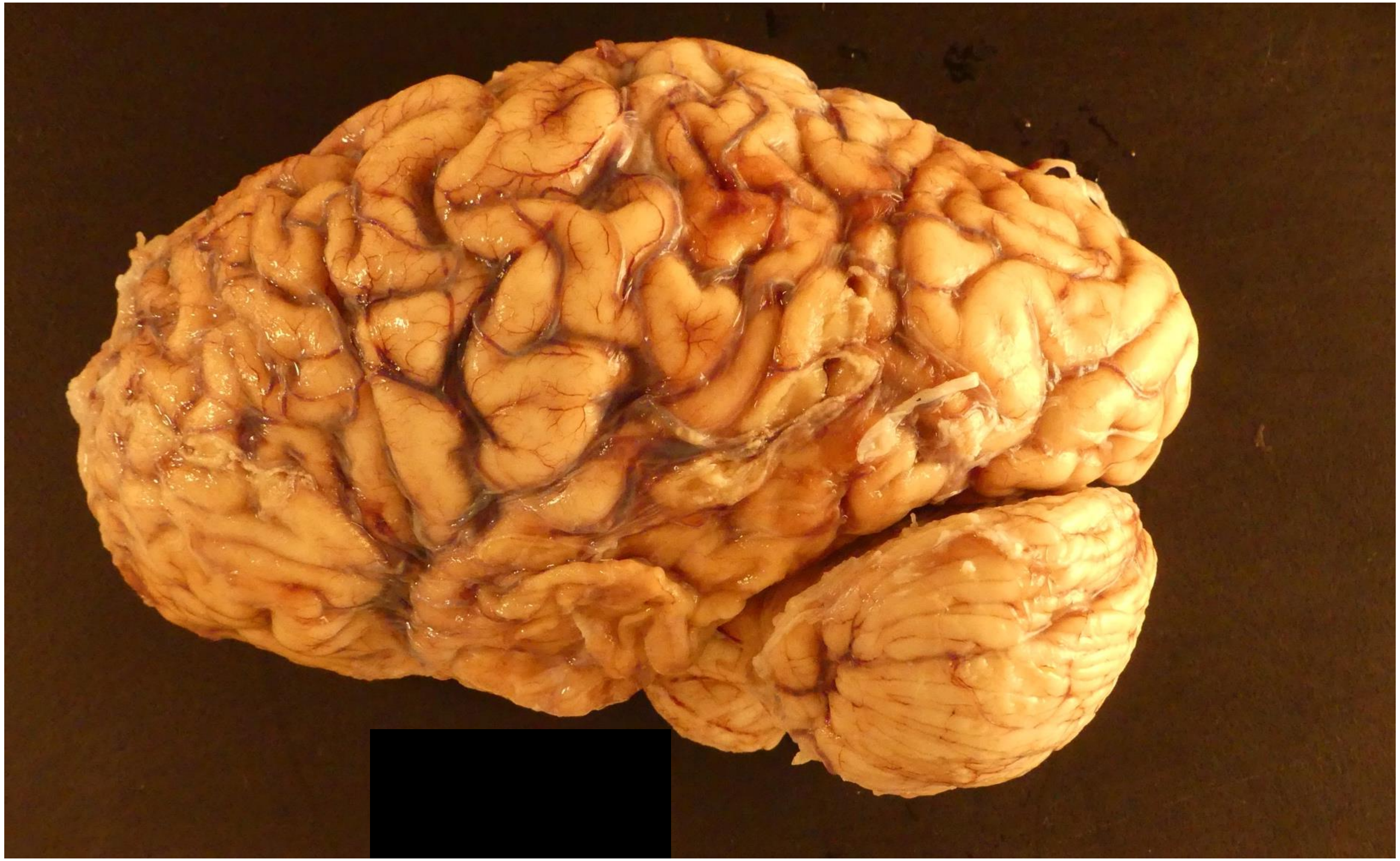

**Supplemental Figure 2:** Graphs of age vs. hippocampal/cortical tau ratio are shown for all AD clusters. Similarly to Figure 2, data points from clusters are shown in red against all other data points from AD cases.

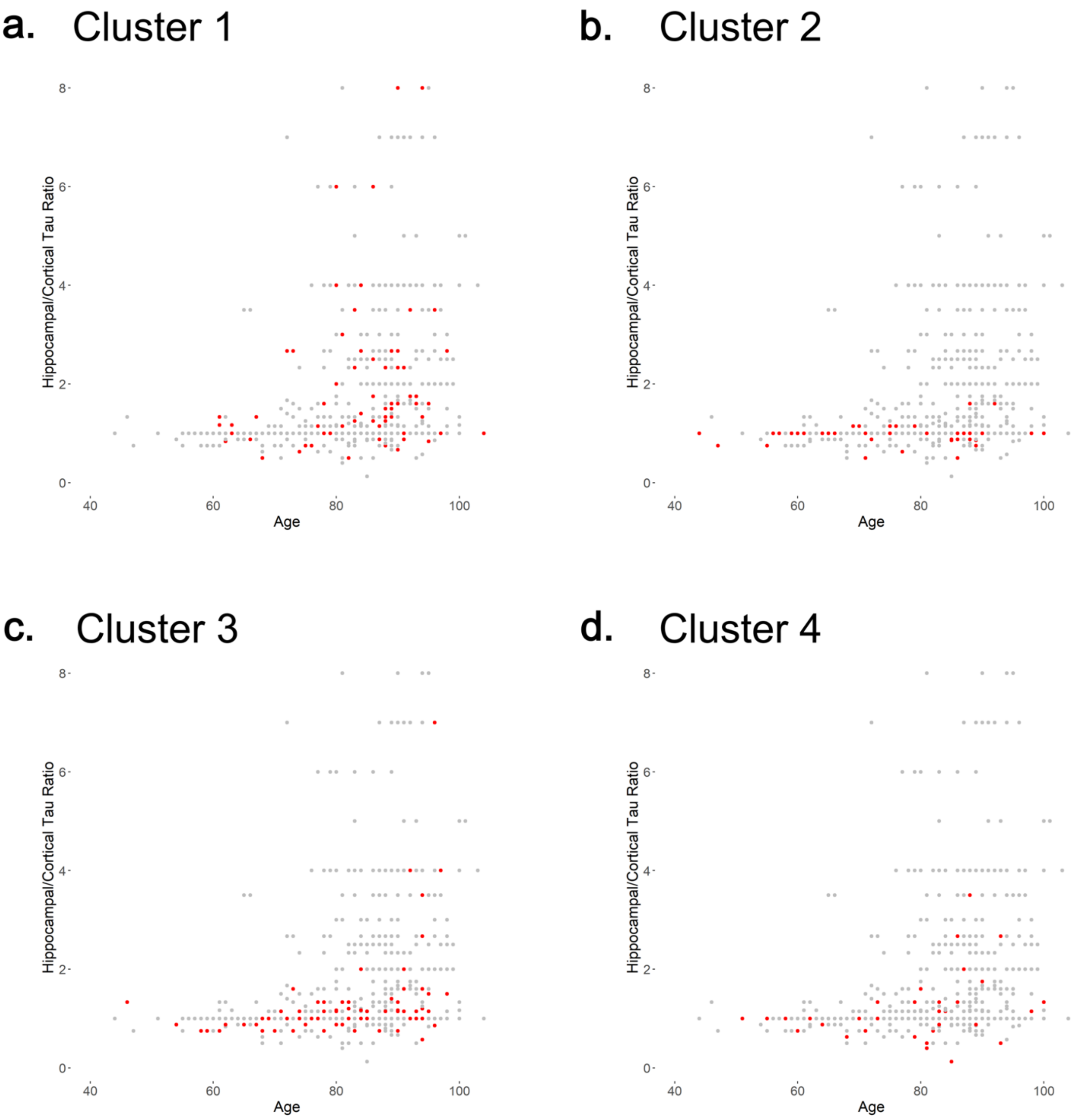

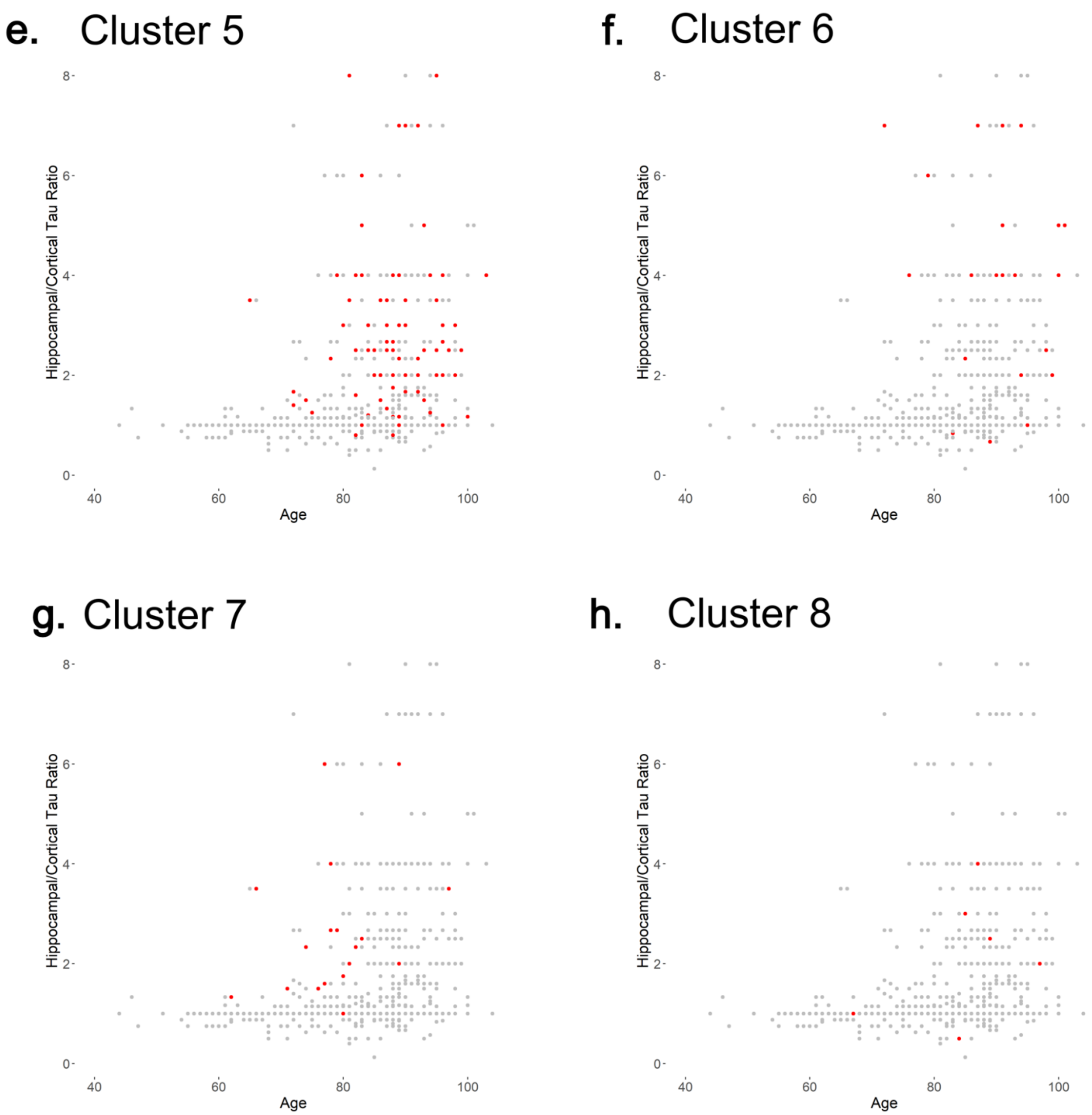

e. Cluster 5
f. Cluster 6
g. Cluster 7
h. Cluster 8
Hippocampal/Cortical Tau Ratio
Age
0
2
4
6
8
40
60
80
100

**<u>Supplemental Figure 3:</u>** Tau (NPT and NFT) and LB correlation charts, incorporating all available NFT and NPT values (Figure 4, panels B and C are a subset of these panels). Cortical tau variables continue to dominate the significant associations with LB variables for the cluster 4 and 7 analysis, even when all NFT and NPT variables are included. Starred boxes = FDR-adjusted p-value < 0.05.

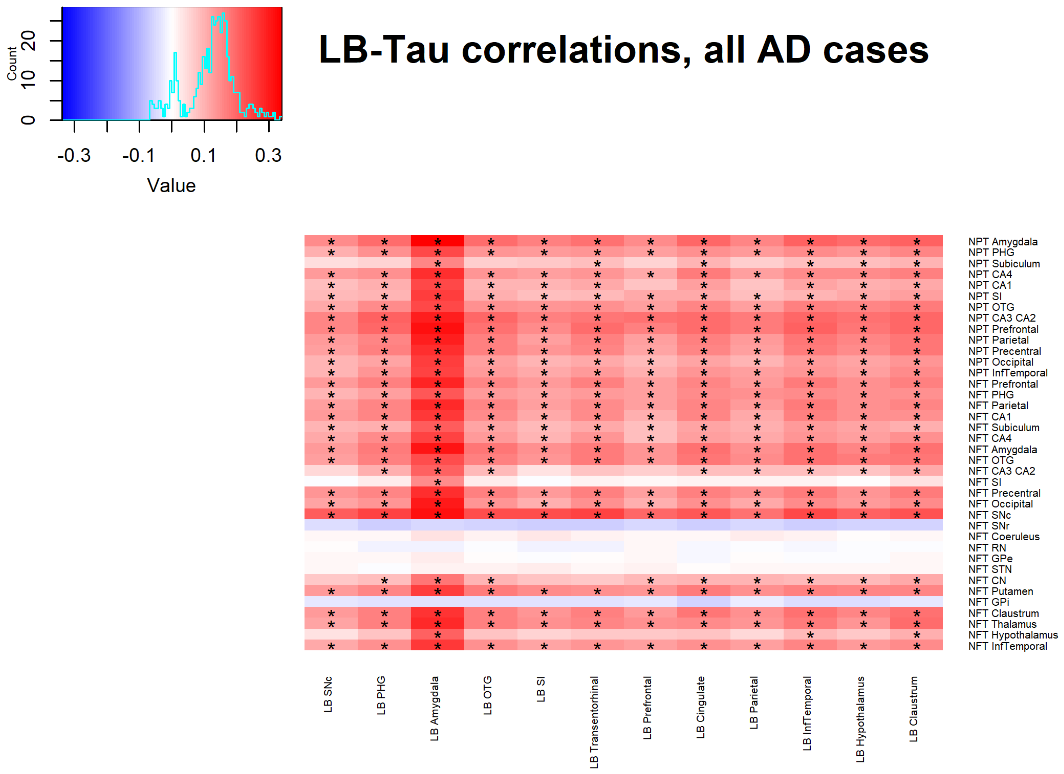


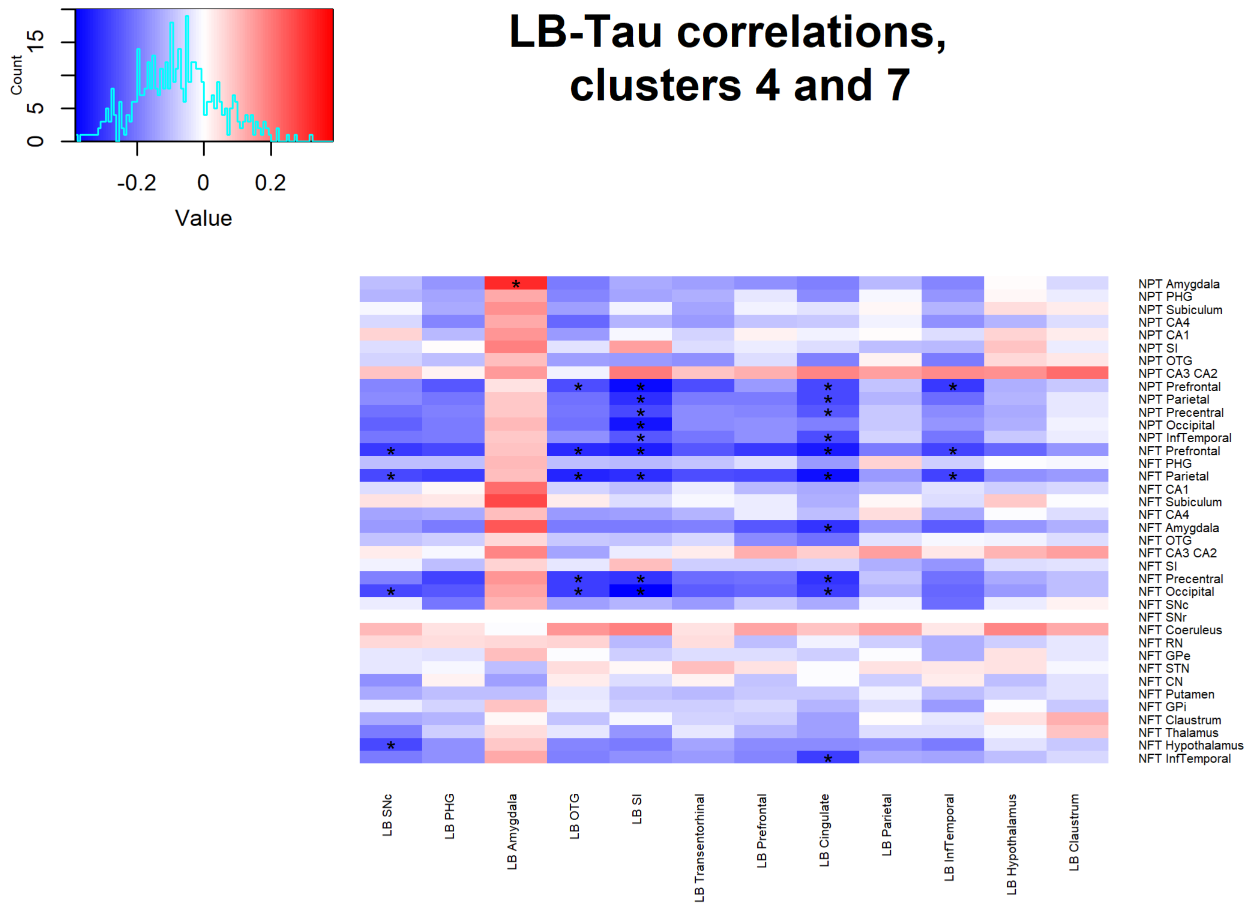
